# Protocol for the Pathways Study: a realist evaluation of staff social ties and communication in the delivery of neonatal care in Kenya

**DOI:** 10.1101/2022.06.27.22276975

**Authors:** C Wanyama, C Blacklock, J Jepkosgei, M English, L Hinton, J McKnight, S Molyneux, M Boga, P Musitia, G Wong

## Abstract

**Introduction:** Initiatives to improve the quality of neonatal care in low- and middle-income countries are vital to meet the Sustainable Development Goal 3 of reducing to at most 12 neonatal deaths per 1,000 by the year 2030. These initiatives have included enhancing human resources for health (HRH), and post-basic neonatal nurse training. However, the informal social ties that health workers form with colleagues and how these influence application of learned knowledge and skills and individual and group behaviours and norms in the workplace, have remained neglected in health systems research.

This study seeks to better understand these relational components in Kenyan neonatal care, and how such understanding might improve design and implementation of quality improvement interventions targeting health workers’ behaviours. The Pathways Study is a realist evaluation which will develop theory for guiding quality improvement interventions.

**Methods and analysis:** We will collect data in two phases. Phase One will be a case study of 2 large urban public hospitals in Kenya, where we will conduct: non-participant observation of hospital staff during patient care and hospital meetings, a social network questionnaire with staff, in-depth interviews, key informant interviews, and focus group discussions. Data will be collected purposively and analysed using a realist logic of analysis, with interim analyses including thematic analysis of qualitative data and quantitative analysis of social network metrics.

Phase Two will be a stakeholder workshop in which findings from Phase One are presented, discussed and theory refined. Recommendations for theory-informed interventions will be developed, to enhance quality improvement efforts in Kenyan hospitals.

**Ethics and dissemination:** Ethical approval for the Pathways Study has been received from Kenya Medical Research Institute (KEMRI/SERU/CGMR-C/241/4374) and Oxford Tropical Research Ethics Committee (OxTREC 519-22). Findings will be shared with the two study hospitals, relevant educational institutions, KEMRI-WELLCOME Trust Research Programme and the University of Oxford. Study findings will also be disseminated in seminars, local and international conferences, and as academic theses and research articles published in open access scientific journals.

**Strengths and limitations of this study:** 

**Strengths:**

- Realist evaluation will enable development of programme theory, which will be useful for informing the practical design and implementation of quality improvement interventions in neonatal units.
- The Pathways Study is the first to use social network analysis to explore the influence of staff social ties on the delivery and quality of neonatal care in Kenya.

**Limitations:**

- Relational ties and other aspects of health systems ‘software’ are notoriously difficult to capture and measure in health systems research.
- The Pathways Study uses a mixed methods approach to collect diverse data, but it is possible that some relevant data may still not be captured. The research team will mitigate this risk by using an iterative and exploratory approach to data collection and analysis, seeking triangulation of emergent findings, promoting reflexivity of the research team, and sense-checking emergent findings with relevant stakeholders, whilst at the same time comparing with substantive sociological theory.

## Introduction

Despite significant reductions in mortality rates across other age groups among children under-5 in recent years, the neonatal mortality rate in Kenya has remained relatively unchanged.(1,2) In response to this disparity, efforts are ongoing to improve the quality of neonatal care, the majority of which is delivered from public health facilities and mostly by nurses.(3) Such improvement efforts have included the recent introduction of new technologies in selected neonatal units, establishing routine neonatal data collection and feedback systems, and ongoing investment in the neonatal workforce through training and workplace support.(4,5)

However, a more detailed understanding of tacit human factors, or health systems ‘software’,(6) that also influence the quality of neonatal care delivered in these hospitals is now needed, to inform better focused design and implementation of improvement efforts (e.g. the relationships between staff, values, norms and the social and cognitive skills that complement clinical technical skills).(7) Indeed, such ‘software’ factors can profoundly influence acquisition of new knowledge and behaviours and norms at individual and group level,(8–11) and thus are determinants of quality care.(12,13) Moreover, individual health worker performance is shaped not only by technical clinical competencies and experiences, but also by social and cognitive skills (cognitive, social and personal resource skills such as communication, team work, leadership, situational awareness, assertiveness and decision making, coping with stress and managing fatigue etc).(14) Relational ties among the workforce and other ‘software’ aspects matter greatly in the delivery of quality neonatal care, for example care pathways, clinical guidelines, knowledge, and other competencies, including those that are transferred from an educational setting such as those gained during post-basic nurse training, to a clinical care environment.(10) Communication which impacts on health worker decision making may furthermore take place in ‘back stage’ ad-hoc opportunistic exchanges, for example in hospital corridors,(8) which can present challenges for capture in research. Despite these challenges for researchers, understanding the ways in which health workers communicate and work with one another is vital. Understanding ‘software’ is particularly important in the neonatal unit, where patients often have multiple complex problems that require care by a multi-professional team who are often working across different units (e.g. maternity, neonatal unit, paediatrics).

Since the care of neonates is highly dependent on teams of staff working together, relational ties and social networks are likely to be central to the adoption of new or better care practices, such as those promoted in existing improvement interventions in Kenyan neonatal units. Understanding how and why communication occurs between health workers will help to unpack the many complex influences on staff behaviours and patient care. Furthermore, detailed understanding of causation and explanatory mechanisms will help to identify areas amenable to intervention that are likely to result in desirable change, perhaps overlooked by more traditional research approaches. Through constructing explanatory programme theory, practical opportunities for improving software in neonatal care will be identified.(15)

In preparation for the Pathways Study, a comprehensive realist synthesis of the literature was undertaken, with included data mostly from high-income settings.(16) Findings from this synthesis were used to help guide the focus of scientific enquiry in the Pathways study, and informed the current protocol and study tools. The realist synthesis sought to answer: how, why, for whom, to what extent and in what contexts, do the social ties of hospital staff influence quality of care. The resultant programme theory comprised of 35 context-mechanism-outcome configurations, organised under four emergent thematic domains: Social group, Hierarchy, Bridging distance and Discourse. The Pathways Study will further develop and refine this provisional programme theory for the specific setting of neonatal units in Kenya, using an open and explorative approach.

The findings of the Pathways Study will make an important contribution to the ongoing work of KEMRI-Wellcome Trust Research Programme researchers in Kenya, particularly work focused on improvement, adoption of technologies and staff communication within the neonatal unit, and by exploring neonatal hospital care beyond the neonatal unit. The new understanding developed during the Pathways Study will be used in the future to better design and target interventions for quality improvement. Findings will also contribute to existing international literature on social networks of the healthcare workforce, the vast majority of which is derived from high income settings.

## Aims and objectives

The aim of the Pathways Study is to explore how, why, for whom and in what circumstances, features of health systems ‘software’ (e.g. values, norms, relationships) between health workers of all cadres caring for neonates in Kenyan hospitals, influence quality of care being targeted by improvement efforts. The specific objectives are:

I. To describe how health workers of all cadres work together to deliver care to newborn babies in Kenyan hospitals, and the kinds of networks, relationships, social ties and personal/team-related socio-cognitive skills (non-technical human skills) that exist within and between the different groups of health workers caring for newborn babies.
II. To examine how these relationships, networks, social ties and related socio-cognitive skills influence the use of nurses’ clinical competencies in newborn care and how these socio-cognitive skills might be utilised to design recommendations to improve neonatal care in Kenyan hospitals.

## Methods and analysis

The Pathways study is a realist evaluation,(15,17) which will employ a mixed methods approach to collect relevant data, which employ a diverse mix of methods to collect relevant data. The data collected will be use to further develop and specify the programme theory from the aforementioned realist review, so that it addresses the research aims. The initial phase of data collection will be from two hospital case study sites, and from a ‘nursing group’ (see below). A stakeholder co-design workshop will be convened following the initial analysis, to further refine theory, and to develop recommendations for practice (Figure 1).

**Figure 1:**
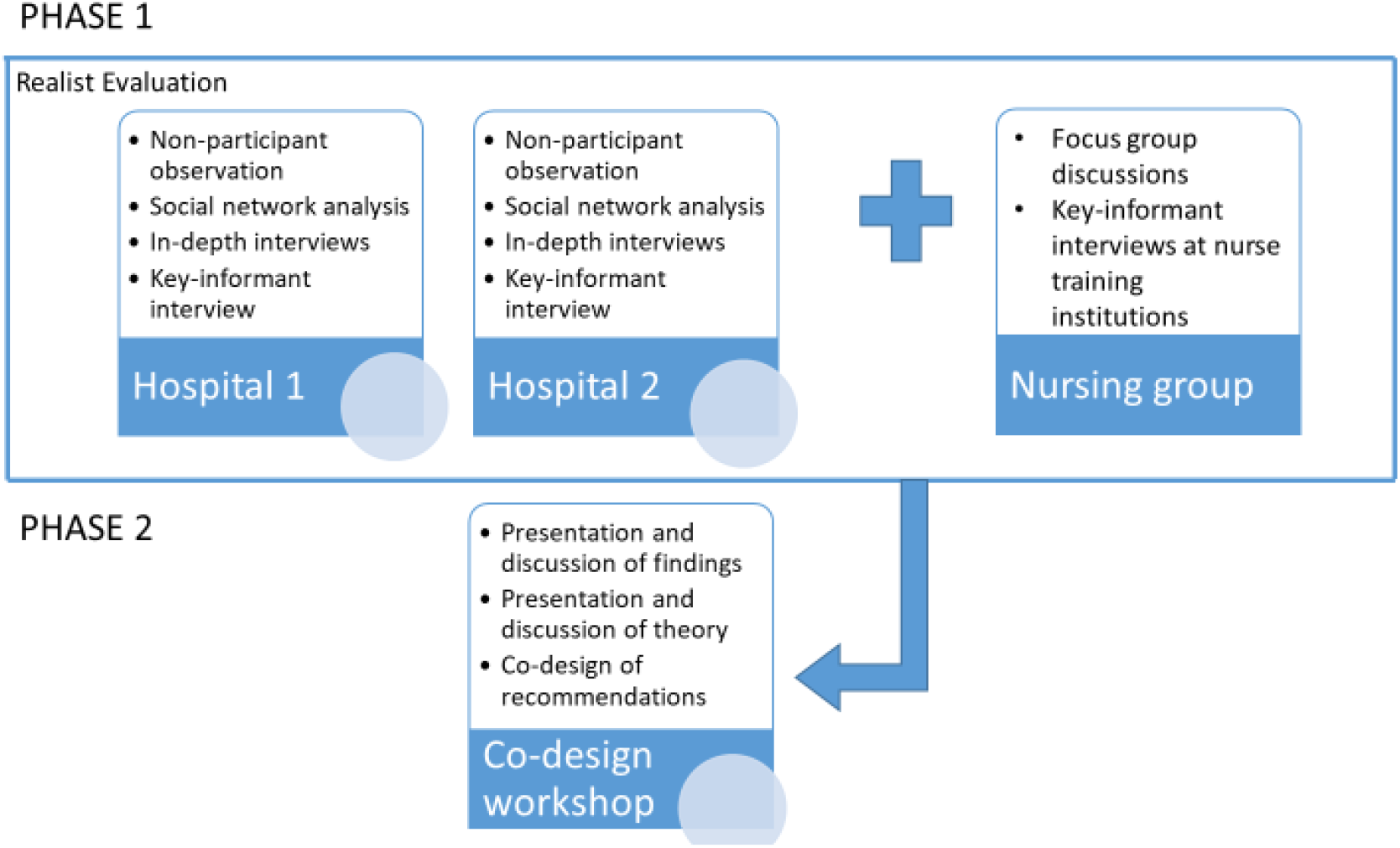
Summary of mixed methods used in the Pathways Study.

## Hospital Case Study Sites

Data will be collected from two large urban public hospitals in Kenya. The study sites have been chosen in part due to ease of access, owing to the existing COVID-19 restrictions to movements. They are also facilities of two contrasts. The Pathways Study will collect data from eligible healthcare workers involved in the provision of care to neonates in the two hospital sites.

Hospital 1 is a large teaching and referral hospital, with a large neonatal unit with a Newborn Intensive Care Unit, and is an implementing site for ongoing initiatives to improve care of neonatal service through the ‘NEST360 (2022 online) Delivering and Sustaining Newborn Technologies’,(5) that seeks to integrate technological solutions within clinical care. The hospital is a training centre for Basic Diploma Nursing and Clinical Medicine students, Bachelor of Science in Nursing and Bachelor of Medicine and Surgery undergraduate students, Post-Graduate Diploma, Higher Diploma and Master’s level training for Paediatric Nursing, Neonatal Nursing, Paediatrics and Child Health and a Fellowship in Neonatology.

Hospital 2 is a large county hospital, with a smaller neonatal unit. Unlike Hospital 1, Hospital 2 is a member of the ‘Clinical Information Network’,(18) which seeks to improve clinical care of children by targeting better use of routine clinical data to inform policy and practice through providing audit and feedback mechanisms and supporting the work of clinical champions in participating network hospitals. Unlike Hospital 1, Hospital 2 is not an implementing site for NEST360(5)

### Nursing group

In addition to the hospital case studies, qualitative data relevant to exploring socio-cognitive skills and the implementation of learned knowledge and skills in the workplace, will be collected from a purposively sampled nursing graduate group and nurse educators from affiliated nurse training institutions.

### Stakeholder group

A group of relevant stakeholders will be invited to participate in a co-design event, following initial data collection and analysis. Co-design outputs will be refined theory and associated recommendations for practice.

Inclusion and exclusion criteria for participation are shown in Table 1.

## Data collection procedures

We will conduct training for all researchers (including research assistants and study clinicians in the various study sites), including the requirements of their roles and how to collect the data using the study data collection tools. Development of the data collection tools (such as observation checklists, interview guides and social network questionnaire) was informed by our previous realist synthesis of the literature.(16) These data collection tools will be piloted amongst the study researchers and colleagues from KEMRI-Wellcome whose work includes some engagements with CIN hospitals, and adapted prior to the start of data collection. To give feedback on the ongoing research process itself, including the piloting and adapting data collection tools, we will also convene a steering group of invited experts.

During data collection and analysis procedures, researchers will adopt a reflexive approach, i.e. all members of the research team will be encouraged to keep aware of their own positionality and biases. Sampling will be purposive, seeking relevant data. The research team will intentionally seek, share and discuss potential biases throughout the course of the Pathways Study, and will mitigate as much as possible through openness and transparent and well documented processes. Training will help to improve individual self-awareness of researchers. The Kenyan research team, consisting of the Principal Investigator and an assistant research officer will meet regularly to compare and discuss emerging findings.

Data collection methods are listed below, with additional details provided in Table 2.

### Non-participant observation

In the two hospital case study sites, we will observe health workers in the neonatal care units/departments they are working. Sampling of shifts (6 per hospital) will be discussed with the unit manager and shifts will be purposively chosen to select a range of different shifts, and to capture variation. A typical shift varies from hospital to hospital, but ranges from 6–8-hour day shift to 12-14 hour night shift. A sample of relevant hospital meetings will also be observed at each hospital site. Consent will be sought from staff at the start of the shift, and observations by the researcher will continue throughout the entire shift to try to mitigate any possible changes in staff behaviour due to being observed. Six shifts will be observed per hospital, to also try to mitigate any potential changes in staff behaviour due to being observed. Though not the focus of observations, caregivers will also have the presence of the researcher explained to them and be given the option to opt out of them/their baby being present in observations of staff.

### Social network questionnaire

We will invite all health workers to participate (a complete network) who are involved in delivering neonatal care from relevant departments (i.e. neonatal unit, paediatric and maternity wards, and the allied departments: laboratory, pharmacy, radiology, nutrition and patient support services). If the number of staff involved in delivering neonatal care exceeds 120, those staff who are most directly involved in the delivery of neonatal care by their place of work being on the neonatal unit, paediatric or maternity wards, will be invited to participate in the social network questionnaire in the first instance.

A complete staffing list will be compiled by speaking to the management of the hospital/units and relevant student representatives (list will include names of staff of all cadres, students, and all shifts). The researcher will assign a random ID code to each member of staff on the list, using a random sequence generator. Participants will be asked to identify colleagues they would speak to from a staffing list, and the researcher will use a separate research list to locate a corresponding ID code for each member of staff/student named during the social network questionnaire. Demographic data will be collected to identify and better understand homophily (those with similar demographics forming preferential ties with one another) within the network, and where it occurs.(9) This will help to understand for example, whether cadre/gender/academic training is associated with connections in the workplace. If conducted online, participants will be asked to name colleagues without using the roster prompt, to avoid electronic sharing of staff lists.

### In-depth interviews

We will conduct purposive sampling to obtain data from approximately 35 study participants from each of the participating hospitals (total of approximately 70). We will seek variation of interview participants to identify and explore diversity of views and experience, for example variation in cadres, units, and demographics.

### Key Informant Interviews

One representative from management will be invited to participate in a key informant interview, from each of the two hospitals and from associated training institutions.

### Focus group discussions

Multi-stage sampling (convenience and snowballing then stratified sampling) will be used for focus group discussions of basic diploma and/or degree nursing graduates, specialist paediatric/neonatal nursing graduates and nursing educators. Participants from training institutions (nursing educators) will be recruited purposively. For nurse graduates, informal/formal professional nursing graduate social media group fora, such as WhatsApp and Telegram, will be used to invite individuals to participate in the study, followed by snowballing to reach those who are not on the social media platforms. This will be followed by stratification (1:1:1) to represent the various professional cadres, training programmes and facilities where they practice.

### Stakeholder Workshop

We will conduct purposive sampling to obtain data from at least one representative amongst the various stakeholders, to achieve a sample size of 10-15.

## Analyses

The research objectives of the Pathways Study which aim to build theory, and the mixed methods approach to data collection, make the use of a realist logic of analysis an appropriate scientific approach for analysis and interpretation of data.

A realist programme theory will be developed, and recommendations for practice will be made from the developed theory. An overaching realist programme theory will be developed through examination of data, initially by seeking out semi-regular patterns of outcomes that occur within the emerging programme theory and then developing causal explanations for these outcomes in the form of context-mechanism-outcome configurations (CMOCs).(19) The initial programme theory (and its consitituent CMOCs) for the Pathways Study will be based on that from our existing realist review.(16) The data collected will be analysed using the same realist logic of analysis to further develop the programme theory. Hence, the programme theory from the realist review will provide initial causal explanations that can be ‘tested’ (confirmed, refuted or refined) against the primary data collected for the realist evalaution.(15) Where indicated by our interpretations of the data, additional CMOCs will be added to the initial programme theory – thus further developing the understanding captured in the emerging programme theory.

Prior to the application of the realist logic of analysis we will do some additional preparatory data analysis, for example thematically analysing the qualitative data and using social network approaches to analyse the quantitative data:

- Qualitative data from the study collected during non-participant observations, in-depth interviews and focus groups, and the stakeholder co-design event will initially be analysed thematically using NVivo software. This will lead to inductively developing a theoretical framework. Qualitative data on socio-cognitive aspects of the study (key informant interviews and in-depth interviews) will be iteratively analysed using a grounded theory approach following an Open-coding →Axial□-coding _→_Selective-coding approach to develop an explanatory theoretical framework on the role of socio-cognitive skills in the utilization of clinical competencies by nurses.
- Quantitative data collected from a Social Network Questionnaire will be analysed with standard SNA metrics (e.g. density, centrality, etc.) using Gephi free software and/or R to develop sociograms and a set of network measures.

Following the initial phase of realist anlysis and theory building, a group of stakeholders will be invited to review and further refine the theory, based on their expert knowledge and experiences. Through engagement with these stakeholders, explanatory theory will be further refined, and used to develop a set of recommendations for practice.

## Patient and Public Involvement

The Pathways Study research team includes a representative from each of the two hospital study sites and training institutions as study collaborators. The role of study site collaborators is to provide useful linkages for facility engagements before, during and after the study as well as facilitate the smooth conduct of the research by contributing to data collection through arranging for meetings with study participants. During the study, an advisory steering group will be formed to review study findings and provide useful comments and expert advisory on study data analysis and interpretation of the study findings.

## Supporting information

Information sheets and consent forms

## Data Availability

All data produced in the present study are available upon reasonable request to the authors

## Ethics and dissemination

Ethical approval for the Pathways Study has been received from Kenya Medical Research Institute (KEMRI/SERU/CGMR-C/241/4374) and Oxford Tropical Research Ethics Committee (OxTREC 519-22).

Insights from the Pathways Study will add new understanding and a different theoretical lens to ongoing work by KEMRI-Wellcome Trust Research Programme in Kenya, particularly work focused on health workforce communication in neonatal care. Findings will also provide a useful addition to the existing body of literature on social networks of the hospital workforce, which is almost entirely derived from high income settings. The explanatory power of realist programme theory in specifically developing new understanding around the chains of causation associated with social ties of hospital staff, will be useful for designing and targeting interventions to enhance how such ties might positively contribute to change efforts in the neonatal unit. In addition, social network analysis (SNA) is a relatively new methodology in healthcare research, and so the practical learning gained during the research process will be shared with KEMRI-Wellcome Trust Research Programme colleagues and others and will be useful for those intending to use SNA in future research.

Findings will further open-up the window of socio-cognitive skills to policymakers and other stakeholders in Kenya, their role in service delivery quality and by way of interventions influence health workforce socio-cognitive skills strengthening agendas. Findings relating to the role of non-technical socio-cognitive skills in the use of learned clinical competencies in neonatal care, will be used to develop recommendations for nurse educational interventions. Recommendations are likely to include how and when these skills can be incorporated into basic and specialised nursing programmes and in-service continuous professional development training activities. We anticipate that these findings might also be applicable to wider healthcare training programmes.

Findings will be shared with the two study hospitals, relevant educational institutions, and KEMRI-WELLCOME Trust Research Programme and the University of Oxford. Study findings will also be disseminated in seminars, local and international conferences, and as academic theses and research articles published in open access scientific journals.

## Authors’ contributions

This protocol was co-developed by Claire Blacklock (CB) and Conrad Wanyama (CW) as joint authors/co-principal investigators. Mike English (ME), Peris Musitia (PM), Mwanamvua Boga (MB), Geoff Wong (GW), Lisa Hinton (LH), Sassy Molyneux (SM) and Jacob McKnight (JM)contributed to the development of the protocol, and Juliet Jepkosgei (JJ) contributed to the refining of the protocol and study tools for implementation. We received protocol review feedback from Michuki Maina (MM), Betty Kalama (BK) and Naomi Muinga (NM), who were nominated as KEMRI-Coast Centre Committee (CCC) and Alex Hinga and Nancy Kagwanja as KEMRI Centre Scientific Committee reviewers. Both the CCC and CSC review feedbacks were incorporated in the final protocol.

## Funding statement

The Pathways Study is supported by the Wellcome Trust (#207522) through an award to ME as a Senior Fellowship.

CB is studying for a DPhil - this work is supported by the Nuffield Department of Medicine, University of Oxford and the Medical Research Council [grant number MR/N013468/1]. The funders had no role in the preparation of this report or the decision to submit for publication.

## Open Access Policy

*This research is funded in whole or in part by the Wellcome Trust (#207522)*. For the purpose of Open Access, the author has applied a CC-BY public copyright licence to any author accepted manuscript version arising from this submission

## Competing interests statement

All authors declare no competing or conflict of interests.

## Notes

### Competing Interest Statement

The authors have declared no competing interest.

### Funding Statement

The work will be supported in part by Wellcome Trust (#207522) through an award to Mike English as a Senior Fellowship.
Claire Blacklock is studying for a DPhil - this work is supported by the Nuffield Department of Medicine, University of Oxford and the Medical Research Council [grant number MR/N013468/1].

### Author Declarations

The Scientific and Ethics Review Unit of Kenya Medical Research Institute (REF 241/4379) and University of Oxford Tropical Research Ethics Committee (REF 519-22) gave ethical approval for this work

