## Supplementary material for "Protocol for the Pathways Study: a realist evaluation of staff social ties and communication in the delivery of neonatal care in Kenya": Information sheets and consent forms

### Appendix I – KEMRI-Wellcome Trust Research Programme: Participant Information Sheet, patient information poster and verbal consent/permission form for non-participant observation (clinical environments) for: -

**Pathways Study Form Ia: Non-participant observation (for individuals working in neonatal care)**

**Study title: Pathways Study:**

**Understanding health worker social ties and communication, and their influence in neonatal care.**

**Lay Title: Understanding how health worker interactions influence the care of newborn babies.**

| Institution | Investigators |
| --- | --- |
| KEMRI Wellcome Trust Research Programme | Conrad Wanyama, Prof. Mike English, Dr. Jalemba Aluvaala, Prof Grace Irimu, Joyline Jepkosgei, Peris Musitia, Dr. Dorothy Oluoch, Prof Sassy Molyneux |
| Nuffield Department of Medicine, University of Oxford, UK | Dr Claire Blacklock Dr. Jacob McKnight*,* |
| THIS Institute, University of Cambridge | Prof. Lisa Hinton |
| Nuffield Department of Primary Care Health Sciences, University of Oxford, UK | Dr. Geoff Wong |
| University of Nairobi | Dr. Aluvaala Jalemba, Dr. Joyce Jebet, Prof Grace Irimu |
| Kenyatta National Hospital | Catherine Ngugi, Josephine Bariu |
| Mbagathi County Referral Hospital | Norah Chebet |
| Kenya Medical Training College | Everlyne Abuga |

You are being asked to take part in a study. The box below tells you important things you should think about before deciding to join the study. We will provide more detailed information below the box. Please take time to read the following information carefully and discuss it with others if you wish to and ask questions about any of the information before you decide to participate. You may also wish to talk to your family or friends about this study, before agreeing to join.

**Key Information for You to Consider**

- **Voluntary Consent**. You are being asked to volunteer for a research study. You can choose whether you would like to participate or not. If you do agree you can change your mind at any time and withdraw from the research. This will not affect your work now or in the future.
- **Purpose**. We are doing this research to better understand how health workers and teams work together to deliver neonatal care, including how changes might impact on different people in the team and how communication might be improved. The research will be used to inform how quality improvement interventions are implemented in hospitals. You have been invited to participate in this research because you work in neonatal care, or you work in the hospital and support neonatal care in some way.
- **Duration.** This observation will last 6-8 hours of the entire shift
- **Procedures and Activities.** We will be observing more generally how staff are working together in this unit during working hours. We will also observe a sample of hospital meetings. Researchers will take field notes during observations.

**Who is carrying out this study and what is this study about?**

- This study is being carried out by KEMRI in collaboration with the University of Oxford, UK. KEMRI is a Kenyan government organization that carries out medical research to find better ways of preventing and treating illness in the future for everybody’s benefit.
- The study will involve observing how people interact with each other while at workplace (who they talk to, what they talk about and how they form interactions and connections). We will also ask health providers what they know, feel or do about these interactions and how these interactions impact care of neonates.
- We want to better understand how health workers and teams work together to deliver neonatal care, including how changes might impact on different people in the team and how communication might be improved. We will do this by observing these interactions during all working shifts. We will then ask about whom health workers are likely to seek advice from when faced with a difficulty (and why). We will also observe a sample of hospital meetings.

**Why do you want to talk to me and what does it involve?**

- We have selected observing people while at work as this is the best way to understand social interactions and communications with each other. You will be observed because the environment you work either directly provides or supports the provision of neonatal care in this hospital. Because this is a non-participant observation, we will not be involved in the care you give to neonates. Researchers will take written field notes. No events will be audio or video-recorded.

**Are there any risks or disadvantages to me for taking part?**

- There are no risks for taking part in the study. However, questions around interactions and social connections can be sensitive as they may include to a discussion about difficulty relationships at workplace.

The observation will take 6-8 hours during the entire shift But we will aim to have only minimal interruption to your work

**Are there any advantages to me for taking part?**

- There are no individual benefits to taking part in this study. In participating in this study, you will contribute to knowledge of social connections and ties, including knowledge of factors that facilitate or hinder use of knowledge and skills acquired during trainings. This finding may help in making recommendations in the design of interventions that improve quality of care of neonates in Kenya and elsewhere in the future, for example through developing new health policies.

**Data Protection**

**Who will have access to information about me in this research?**

- All our research records are stored securely in locked cabinets and password protected computers and are accessed only by authorized persons.
- The personally identifiable information you provide will be used only for study purposes such as contact you for scheduling of visits and verification of consent through signed consent forms.
- We will share anonymized individual and summary information we collect or generate with our partner institution, University of Oxford in ways that do not reveal individual participants’ identities.
- Anonymised data will be stored in a secure KEMRI-Wellcome Trust Research Programme repository and may be used for future research.
- KEMRI will keep any keep any personally identifiable information about you from this study for 10 years after the study has finished in accordance with applicable Data Protection Requirements both in Kenya and in the UK. You have the right to access the personal data we hold that pertains to you, to object to or make corrections to the processing of all or part of the personal data. However, this might be limited due to coding which might make unblinding difficult.

**Will the research be published? Could I be identified from any publications or other research outputs?**

- The research will be published. We might however include direct quotations, but without identifying you, in any research outputs.
- In future, information collected or generated during this study may be used to support new research by other researchers in Kenya or other countries on improving newborn service delivery. In all cases, we will only share information with other researchers in ways that do not reveal individual participants’ identities. For example, we will remove such as their names and where they live and replace this information with number codes. Any future research using information from this study must first be approved by a local or national expert committee to make sure that the interests of participants and their communities are protected.
- University of Oxford is responsible for ensuring that staff involved in the study in Kenya adhere to the safe and proper use of any personal information you provide. You can contact the research team for any further information about how your data will be managed. Further information about your rights with respect to your personal data is available from <https://compliance.admin.ox.c.uk/individual-rights>

**Who has allowed this research to take place?**

All research at KEMRI has to be approved before it begins by an independent national committee, an international committee and a committee at the KEMRI Wellcome Trust Research Programme. The committees look carefully at planned work before it begins, and must agree that the research is important, relevant to Kenya and follows nationally and internationally agreed research guidelines. This includes ensuring that all participants’ safety and rights are respected. This study will also seek ethics approval from a subcommittee of the University of Oxford Central University Research Ethics Committee.

**What will happen if I refuse to participate?**

All participation in research is voluntary. You are free to decide if you want to take part or not. If you do agree you can change your mind at any time without any consequences. You can withdraw yourself from the study, without giving a reason, and without negative consequences, by advising us of this decision. The deadline by which you can withdraw from the study is September 2022, because we expect that data from this study would have already began being analysed by this time. If you withdraw from the study after this date, all information that you have already provided will still be included in the analysis. If you withdraw from the study before this date, information you have already provided may still be included if analysis has already occurred, however the information you have provided will not be used as quotes and the information will be removed from any data repositories.

**What if I have any questions?**

You are free to ask me any question about this research. If you have any further questions about the study, you are free to contact the research team using the contacts below:

Mr. Conrad Wanyama, KEMRI Wellcome Trust Research Programme, P.O. Box 230, Kilifi. Telephone: [mobile 0721275262] or 0700274694

**If you want to ask someone independent anything about this research please contact:**

Community Liaison Manager, KEMRI Wellcome Trust Research Programme, P.O. Box 230, Kilifi. Telephone: 041 7522 063, Mobile 0723 342 780 or 0705 154 386

***And***

### KEMRI-Wellcome Trust Research Programme Information Poster for Pathways Study

**Formal title**: Understanding health worker social ties and communication, and their influence in neonatal care.

**Lay title:** Understanding how health worker interactions influence the care of newborn babies.

**Appendix Ib**: Information Poster

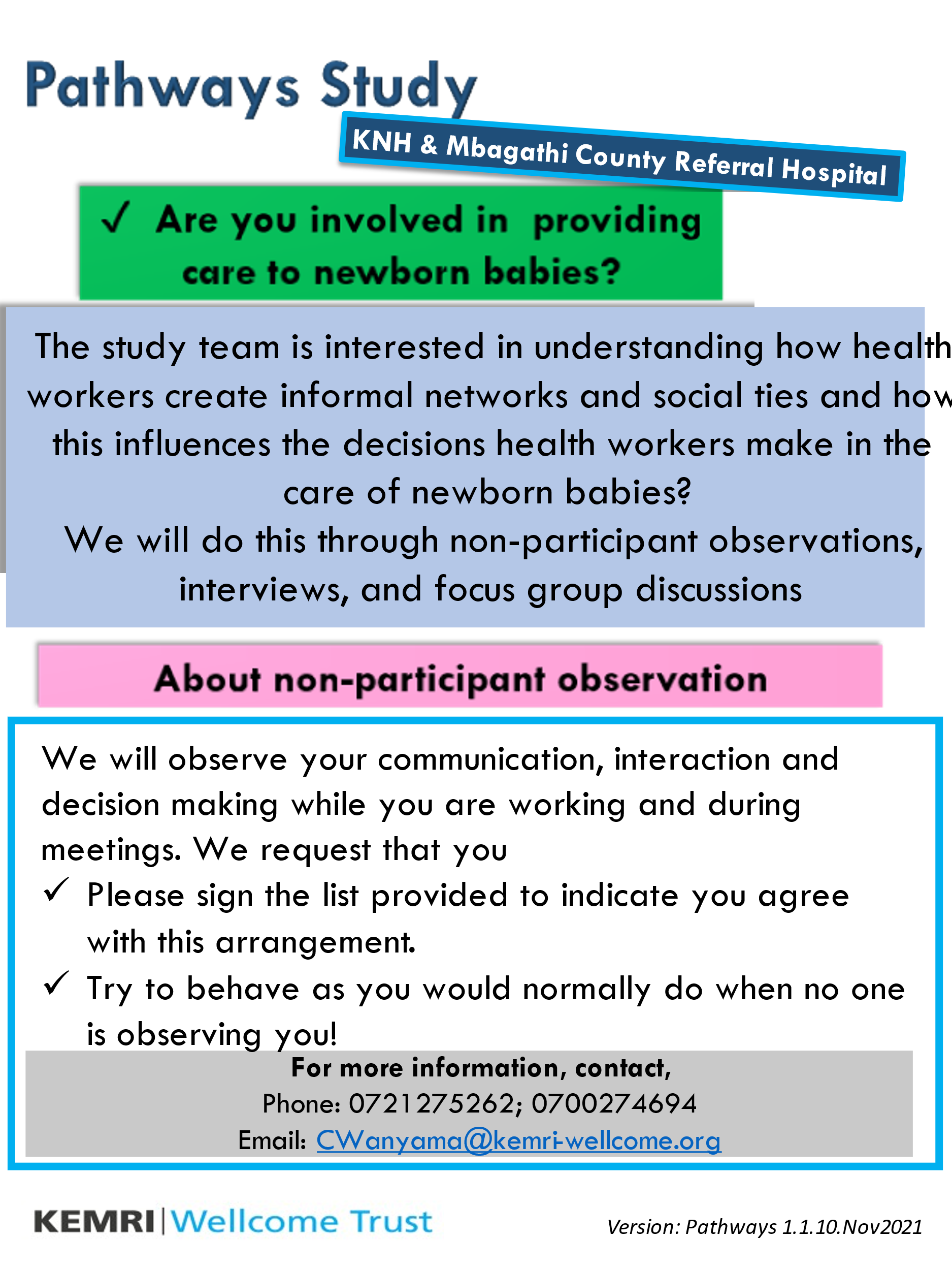

### KEMRI-Wellcome Trust Research Programme: verbal consent/permission form for non-participant observation for staff for: -

**Study title: Pathways Study: Understanding health worker social ties and communication, and their influence in neonatal care.**

**Lay Title: Understanding how health worker interactions influence the care of newborn babies.**

I confirm that I have provided information in the form of a study information poster in distinct areas within the hospital. I also confirm that I have explained the study and the process of observation to the participant(s) named below in accordance with the SOP and that I helped them understand the study and the purpose of the non-participant observation and gave them the chance to ask questions.

I also confirm that I explained to the participant(s) named below that their work will not be audio or video-recorded and that all the aspects of care, interactions and social ties will be used for the research purpose only.

**I confirm that the study participant (s) listed below were present during the briefing session.**

**Shift: ______________________________________________________________________________________**

Present	Cadre code

Participant Name: _______________________________________________________	□

Participant Name: _______________________________________________________	□

Participant Name: _______________________________________________________	□

Participant Name: _______________________________________________________	□

Participant Name: _______________________________________________________	□

Participant Name: _______________________________________________________	□

Participant Name: _______________________________________________________	□

Participant Name: _______________________________________________________	□

Participant Name: _______________________________________________________	□

Participant Name: _______________________________________________________	□

Participant Name: _______________________________________________________	□

Participant Name: _______________________________________________________	□

Participant Name: _______________________________________________________	□

Participant Name: _______________________________________________________	□

Participant Name: _______________________________________________________	□

Participant Name: _______________________________________________________	□

Participant Name: _______________________________________________________	□

Participant Name: _______________________________________________________	□

Participant Name: _______________________________________________________	□

Participant Name: _______________________________________________________	□

Participant Name: _______________________________________________________	□

Participant Name: _______________________________________________________	□

Participant Name: _______________________________________________________	□

Participant Name: _______________________________________________________	□

**Designee/investigator’s signature:** ____________________________ **Date** ____________

**Designee/investigator’s name:** _____________________________**Time** ____________

### Appendix J– KEMRI-Wellcome Trust Research Programme: Participant Information Sheet and Consent form for non-participant observation (hospital meetings) for: -

**Pathways Study Form J: Non-participant observation (for individuals attending hospital meetings)**

**Study title: Pathways Study: Understanding health worker social ties and communication, and their influence in neonatal care.**

**Lay Title: Understanding how health worker interactions influence the care of newborn babies.**

| Institution | Investigators |
| --- | --- |
| KEMRI Wellcome Trust Research Programme | Conrad Wanyama, Prof. Mike English, Dr. Jalemba Aluvaala, Prof Grace Irimu, Joyline Jepkosgei, Peris Musitia, Dr. Dorothy Oluoch, Prof Sassy Molyneux |
| Nuffield Department of Medicine, University of Oxford, UK | Dr Claire Blacklock Dr. Jacob McKnight*,* |
| THIS Institute, University of Cambridge | Prof. Lisa Hinton |
| Nuffield Department of Primary Care Health Sciences, University of Oxford, UK | Dr. Geoff Wong |
| University of Nairobi | Dr. Aluvaala Jalemba, Dr. Joyce Jebet, Prof Grace Irimu |
| Kenyatta National Hospital | Catherine Ngugi, Josephine Bariu |
| Mbagathi County Referral Hospital | Norah Chebet |
| Kenya Medical Training College | Everlyne Abuga |

You are being asked to take part in a study. The box below tells you important things you should think about before deciding to join the study. We will provide more detailed information below the box. Please take time to read the following information carefully and discuss it with others if you wish to and ask questions about any of the information before you decide to participate. You may also wish to talk to your family or friends about this study, before agreeing to join.

**Key Information for You to Consider**

- **Voluntary Consent**. You are being asked to volunteer for a research study. You can choose whether you would like to participate or not. If you do agree you can change your mind at any time and withdraw from the research. This will not affect your work now or in the future.
- **Purpose**. We are doing this research to better understand how health workers and teams work together to deliver neonatal care, including how changes might impact on different people in the team and how communication might be improved. The research will be used to inform how quality improvement interventions are implemented in hospitals.
- **Duration.** Your participation in this study will last for the entire period of this meeting all the way until everyone has dispersed.
- **Procedures and Activities.** We will be observing more generally how staff are communicating and interacting (verbally and non-verbally) during the formal meeting and shortly before/after the meeting (who they talk to, what they talk about and how they form interactions and connections). We will also ask attendees what they know, feel, or do about these interactions and how these interactions impact care of neonates

**Who is carrying out this study and what is this study about?**

- This study is being carried out by KEMRI in collaboration with the University of Oxford, UK. KEMRI is a Kenyan government organization that carries out medical research to find better ways of preventing and treating illness in the future for everybody’s benefit.
- We want to better understand how health workers and teams work together to deliver neonatal care, including how changes might impact on different people in the team and how communication might be improved. We will do this by observing these interactions during meetings. We will all adhere to the requisite Ministry of Health COVID-19 protocols.

**Why do you want to talk to me and what does it involve?**

We have selected observing meetings as they shape the direction of how people interact and work together in the care of patients. Because this is a non-participant observation, we will not interrupt the discussions nor make any contributions (unless if want us to). The meetings will not be audio or video recorded.

**Are there any risks or disadvantages to me for taking part?**

There are no risks for taking part in the study. However, an external attendee in hospital meetings might create some discomfort when sensitive internal matters are being discussed. We will aim to maintain confidentiality of all matters discussed in the meeting, and only focus on the purpose of the research.

**Are there any advantages to me for taking part?**

There are no individual benefits to taking part in this study. In participating in this study, you will contribute to knowledge of social connections and ties, including knowledge of factors that facilitate or hinder use of knowledge and skills acquired during nurse trainings. This finding may help in making recommendations in the design of interventions that improve quality of care of neonates in Kenya and elsewhere in the future, for example through developing new health policies.

**Data Protection**

**Who will have access to information about me in this research?**

- All our research records are stored securely in locked cabinets and password protected computers and are accessed only by authorized persons.
- The personally identifiable information you provide will be used only for study purposes such as contact you for scheduling of visits and verification of consent through signed consent forms.
- We will share anonymized individual and summary information we collect or generate with our partner institution, University of Oxford in ways that do not reveal individual participants’ identities.
- Anonymised data will be stored in a secure KEMRI-Wellcome Trust Research Programme repository and may be used for future research.
- KEMRI will keep any keep any personally identifiable information about you from this study for 10 years after the study has finished in accordance with applicable Data Protection Requirements both in Kenya and in the UK. You have the right to access the personal data we hold that pertains to you, to object to or make corrections to the processing of all or part of the personal data. However, this might be limited due to coding which might make unblinding difficult.

**Will the research be published? Could I be identified from any publications or other research outputs?**

- The research will be published. We might however include direct quotations, but without identifying you, in any research outputs.
- In future, information collected or generated during this study may be used to support new research by other researchers in Kenya or other countries on improving newborn service delivery. In all cases, we will only share information with other researchers in ways that do not reveal individual participants’ identities. For example, we will remove such as their names and where they live and replace this information with number codes. Any future research using information from this study must first be approved by a local or national expert committee to make sure that the interests of participants and their communities are protected.
- University of Oxford is responsible for ensuring that staff involved in the study in Kenya adhere to the safe and proper use of any personal information you provide. You can contact the research team for any further information about how your data will be managed. Further information about your rights with respect to your personal data is available from <https://compliance.admin.ox.c.uk/individual-rights>

**Who has allowed this research to take place?**

All research at KEMRI has to be approved before it begins by an independent national committee, an international committee and a committee at the KEMRI Wellcome Trust Research Programme. The committees look carefully at planned work before it begins, and must agree that the research is important, relevant to Kenya and follows nationally and internationally agreed research guidelines. This includes ensuring that all participants’ safety and rights are respected. This study will also seek ethics approval from a subcommittee of the University of Oxford Central University Research Ethics Committee.

**What will happen if I refuse to participate?**

All participation in research is voluntary. You are free to decide if you want to take part or not. If you do agree you can change your mind at any time without any consequences. You can withdraw yourself from the study, without giving a reason, and without negative consequences, by advising us of this decision. The deadline by which you can withdraw from the study is September 2022, because we expect that data from this study would have already began being analysed by this time. If you withdraw from the study after this date, all information that you have already provided will still be included in the analysis. If you withdraw from the study before this date, information you have already provided may still be included if analysis has already occurred, however the information you have provided will not be used as quotes and the information will be removed from any data repositories.

**What if I have any questions?**

You are free to ask me any question about this research. If you have any further questions about the study, you are free to contact the research team using the contacts below:

Mr. Conrad Wanyama, KEMRI Wellcome Trust Research Programme, P.O. Box 230, Kilifi. Telephone: [mobile 0721275262] or 0700274694

**If you want to ask someone independent anything about this research please contact:**

Community Liaison Manager, KEMRI Wellcome Trust Research Programme, P.O. Box 230, Kilifi. Telephone: 041 7522 063, Mobile 0723 342 780 or 0705 154 386

***And***

### KEMRI-Wellcome Trust Research Programme: verbal consent/permission form for non-participant observation for hospital meetings for: -

**Study title: Pathways Study: Understanding health worker social ties and communication, and their influence in neonatal care.**

**Lay Title: Understanding how health worker interactions influence the care of newborn babies.**

I confirm that I have explained the study and the process of observation to the participant(s) named below in accordance with the SOP and that I helped them understand the study and the purpose of observation and gave them the chance to ask questions.

I also confirm that I explained to the participant(s) named below that they have the right to refuse to be included in observations and the right to withdraw their permission at any point.

**I confirm that verbal consent was provided (or denied) by the participant(s) listed below who agreed (did not agree) to take part in this research.**

Agree	Refused

Participant Name: _______________________________________________________	□	□

Participant Name: _______________________________________________________	□	□

Participant Name: _______________________________________________________	□	□

Participant Name: _______________________________________________________	□	□

Participant Name: _______________________________________________________	□	□

Participant Name: _______________________________________________________	□	□

Participant Name: _______________________________________________________	□	□

Participant Name: _______________________________________________________	□	□

Participant Name: _______________________________________________________	□	□

Participant Name: _______________________________________________________	□	□

Participant Name: _______________________________________________________	□	□

Participant Name: _______________________________________________________	□	□

Participant Name: _______________________________________________________	□	□

Participant Name: _______________________________________________________	□	□

Participant Name: _______________________________________________________	□	□

Participant Name: _______________________________________________________	□	□

Participant Name: _______________________________________________________	□	□

Participant Name: _______________________________________________________	□	□

Participant Name: _______________________________________________________	□	□

Participant Name: _______________________________________________________	□	□

Participant Name: _______________________________________________________	□	□

Participant Name: _______________________________________________________	□	□

Participant Name: _______________________________________________________	□	□

Participant Name: _______________________________________________________	□	□

**Designee/investigator’s signature:** ____________________________ **Date** ____________

**Designee/investigator’s name:** _____________________________**Time** ____________

### Appendix K– KEMRI-Wellcome Trust Research Programme: Participant Information Sheet and Consent form for Social Network Analysis (SNA) questionnaire for: -

**Pathways Study Form K: Social Network Analysis (SNA) Questionnaire (for health workers directly providing or indirectly supporting the provision of neonatal care)**

**Study title: Pathways Study: Understanding health worker social ties and communication, and their influence in neonatal care.**

**Lay Title: Understanding how health worker interactions influence the care of newborn babies.**

| Institution | Investigators |
| --- | --- |
| KEMRI Wellcome Trust Research Programme | Conrad Wanyama, Prof. Mike English, Dr. Jalemba Aluvaala, Prof Grace Irimu, Joyline Jepkosgei, Peris Musitia, Dr. Dorothy Oluoch, Prof Sassy Molyneux |
| Nuffield Department of Medicine, University of Oxford, UK | Dr Claire Blacklock Dr. Jacob McKnight*,* |
| THIS Institute, University of Cambridge | Prof. Lisa Hinton |
| Nuffield Department of Primary Care Health Sciences, University of Oxford, UK | Dr. Geoff Wong |
| University of Nairobi | Dr. Aluvaala Jalemba, Dr. Joyce Jebet, Prof Grace Irimu |
| Kenyatta National Hospital | Catherine Ngugi, Josephine Bariu |
| Mbagathi County Referral Hospital | Norah Chebet |
| Kenya Medical Training College | Everlyne Abuga |

You are being asked to take part in a study. The box below tells you important things you should think about before deciding to join the study. We will provide more detailed information below the box. Please take time to read the following information carefully and discuss it with others if you wish to and ask questions about any of the information before you decide to participate. You may also wish to talk to your family or friends about this study, before agreeing to join.

**Key Information for You to Consider**

- **Voluntary Consent**. You are being asked to volunteer for a research study. You can choose whether you would like to participate or not. If you do agree you can change your mind at any time and withdraw from the research. This will not affect your work now or in the future.
- **Purpose**. We are doing this research to better understand how health workers and teams work together to deliver neonatal care, including how changes might impact on different people in the team and how communication might be improved. The research will be used to inform how quality improvement interventions are implemented in hospitals. You have been invited to participate in this research because you work in a neonatal care unit, or you work in the hospital and support neonatal care in some way (the delivery unit, post-natal unit, neonatal unit and paediatric unit, laboratory, pharmacy, nutrition, patient support, physiotherapy, orthopaedic/plaster care and occupational therapy).
- **Duration.** Your participation in this study will last 30-45 minutes.
- **Procedures and Activities.** We will ask you some questions for you to respond to while in a quiet, private place. These discussions will either be online, via phone or face-to-face. If conducted face-to-face, the requisite Ministry of Health COVID-19 protocols will be adhered to. If you agree to participate the researcher will ask you to sign a consent form before asking any questions. You can stop or pause the Social Network Analysis (SNA) questionnaire interview at any time, by letting the researcher know. We will take notes during the questionnaire interview,

**Who is carrying out this study and what is this study about?**

- This study is being carried out by KEMRI in collaboration with the University of Oxford, UK. KEMRI is a Kenyan government organization that carries out medical research to find better ways of preventing and treating illness in the future for everybody’s benefit.
- The study involves asking health workers what they know, feel or do when they need help or advice from colleagues, using existing social interactions and networks and how these networks and social interactions impact care of neonates.
- We want to better understand how health workers and teams work together to deliver neonatal care, including how changes might impact on different people in the team and how communication might be improved.

**Why do you want to talk to me and what does it involve?**

- *Selection*

This hospital has several clinical environments where neonates receive care, including many supportive departments that contribute to the care of neonates. You have been selected from your department/unit so that we get experiences from individuals that will give us a true picture of the social networks and connections in the hospital, without preference.

- *About the SNA questionnaire*:

I/my colleague would like to ask you a number of questions about your background and the interactions you have with your colleagues at work. If you do not want to answer any of the questions you may say so and the interviewer will move on to the next question.

The researcher will take written notes during the interview, to complete the questionnaire. A researcher may sometimes need to contact you again, to clarify information.

**Are there any risks or disadvantages to me for taking part?**

- There are no risks for taking part in the study. However, we will ask you some questions about your background and interactions with colleagues.
- We will aim to conduct the interview during your tea or lunch break or when you are free while you are on duty to minimize disruption of your duties and obligation to work.
- If you are attending this SNA questionnaire activity while on your off or leave day (and not on a working day/shift), we will reimburse your transport costs at a rate of Ksh. 600 per person or mileage costs at the rate of Ksh.25/km for the face-to-face interview
- Online/phone questionnaire interviews will be reimbursed with a data voucher worth Ksh.250-500.

**Are there any advantages to me for taking part?**

- There are no individual benefits to taking part in this study. In participating in this study, you will contribute to knowledge of social connections and ties, including knowledge of factors that facilitate or hinder use of knowledge and skills acquired during nurse trainings. This finding may help in making recommendations in the design of interventions that improve quality of care of neonates in Kenya and elsewhere in the future, for example through developing new health policies.

**Data Protection**

**Who will have access to information about me in this research?**

- All our research records are stored securely in locked cabinets and password protected computers and are accessed only by authorized persons.
- The personally identifiable information you provide will be used only for study purposes such as contact you for scheduling of visits and verification of consent through signed consent forms.
- We will share anonymized individual and summary information we collect or generate with our partner institution, University of Oxford in ways that do not reveal individual participants’ identities.
- Anonymised data will be stored in a secure KEMRI-Wellcome Trust Research Programme repository and may be used for future research.
- KEMRI will keep any keep any personally identifiable information about you from this study for 10 years after the study has finished in accordance with applicable Data Protection Requirements both in Kenya and in the UK. You have the right to access the personal data we hold that pertains to you, to object to or make corrections to the processing of all or part of the personal data. However, this might be limited due to coding which might make unblinding difficult.

**Will the research be published? Could I be identified from any publications or other research outputs?**

- The research will be published. We might however include direct quotations, but without identifying you, in any research outputs.
- In future, information collected or generated during this study may be used to support new research by other researchers in Kenya or other countries on improving newborn service delivery. In all cases, we will only share information with other researchers in ways that do not reveal individual participants’ identities. For example, we will remove such as their names and where they live and replace this information with number codes. Any future research using information from this study must first be approved by a local or national expert committee to make sure that the interests of participants and their communities are protected.
- University of Oxford is responsible for ensuring that staff involved in the study in Kenya adhere to the safe and proper use of any personal information you provide. You can contact the research team for any further information about how your data will be managed. Further information about your rights with respect to your personal data is available from <https://compliance.admin.ox.c.uk/individual-rights>

**Who has allowed this research to take place?**

All research at KEMRI must be approved before it begins by an independent national committee, an international committee and a committee at the KEMRI Wellcome Trust Research Programme. The committees look carefully at planned work before it begins, and must agree that the research is important, relevant to Kenya and follows nationally and internationally agreed research guidelines. This includes ensuring that all participants’ safety and rights are respected. This study will also seek ethics approval from a subcommittee of the University of Oxford Central University Research Ethics Committee.

**What will happen if I refuse to participate?**

All participation in research is voluntary. You are free to decide if you want to take part or not. If you do agree you can change your mind at any time without any consequences. You can withdraw yourself from the study, without giving a reason, and without negative consequences, by advising us of this decision. The deadline by which you can withdraw from the study is September 2022, because we expect that data from this study would have already began being analysed by this time. If you withdraw from the study after this date, all information that you have already provided will still be included in the analysis. If you withdraw from the study before this date, information you have already provided may still be included if analysis has already occurred, however the information you have provided will not be used as quotes and the information will be removed from any data repositories.

**What if I have any questions?**

You are free to ask me any question about this research. If you have any further questions about the study, you are free to contact the research team using the contacts below:

Mr. Conrad Wanyama, KEMRI Wellcome Trust Research Programme, P.O. Box 230, Kilifi. Telephone: [mobile 0721275262] or 0700274694

**If you want to ask someone independent anything about this research please contact:**

Community Liaison Manager, KEMRI Wellcome Trust Research Programme, P.O. Box 230, Kilifi. Telephone: 041 7522 063, Mobile 0723 342 780 or 0705 154 386

***And***

### KEMRI-Wellcome Trust Research Programme consent form for SNA questionnaire for: -

**Study title: Pathways Study: Understanding health worker social ties and communication, and their influence in neonatal care.**

**Lay Title: Understanding how health worker interactions influence the care of newborn babies.**

I have had the study explained to me. I have understood all that has been read/explained and had my questions answered satisfactorily. And I agree to take part in this research

**Please initial the sentences that reflect your choices, and then sign below:**

I do wish to be notified by investigators in the event of research findings of possible importance to my facility or myself. **Yes	No**

I agree that the study team use the identifier that I have provided (telephone number, country ID number, etc.) to locate me in the future. **Yes	No**

I agree to the use of quotations in research outputs if I am not identifiable **Yes	No**

I agree to my anonymised research data being used, for future research **Yes	No**

I understand that I can change my mind at any stage and it will not affect me in any way.

| **Signature:** |  | **Date:** |
| --- | --- | --- |
| **Participant Name:** |  | **Time:** |
|  | (Please print name) |  |

--------------------------------------------------------------------------------------------------------------------------------------

I have followed the study procedure to obtain consent from the participant. S/he apparently understood the nature and the purpose of the study and consents to the participation in the study. S/he has been given opportunity to ask questions which have been answered satisfactorily.

**Designee/investigator’s signature:** ____________________________ **Date** ____________

**Designee/investigator’s name:** _____________________________**Time** ____________

(Please print name)

*THE PARTICIPANT SHOULD NOW BE GIVEN A SIGNED COPY TO KEEP*

*……………………………………………………………………………………………………………………………………………………………………………*

### Appendix L– KEMRI-Wellcome Trust Research Programme: Participant Information Sheet and Consent form for In-depth interviews for: -

**Pathways Study Form L: In-depth interview (for health workers directly providing or indirectly supporting the provision of neonatal care)**

**Study title: Pathways Study: Understanding health worker social ties and communication, and their influence in neonatal care.**

**Lay Title: Understanding how health worker interactions influence the care of newborn babies.**

| Institution | Investigators |
| --- | --- |
| KEMRI Wellcome Trust Research Programme | Conrad Wanyama, Prof. Mike English, Dr. Jalemba Aluvaala, Prof Grace Irimu, Joyline Jepkosgei, Peris Musitia, Dr. Dorothy Oluoch, Prof Sassy Molyneux |
| Nuffield Department of Medicine, University of Oxford, UK | Dr Claire Blacklock Dr. Jacob McKnight*,* |
| THIS Institute, University of Cambridge | Prof. Lisa Hinton |
| Nuffield Department of Primary Care Health Sciences, University of Oxford, UK | Dr. Geoff Wong |
| University of Nairobi | Dr. Aluvaala Jalemba, Dr. Joyce Jebet, Prof Grace Irimu |
| Kenyatta National Hospital | Catherine Ngugi, Josephine Bariu |
| Mbagathi County Referral Hospital | Norah Chebet |
| Kenya Medical Training College | Everlyne Abuga |

You are being asked to take part in a study. The box below tells you important things you should think about before deciding to join the study. We will provide more detailed information below the box. Please take time to read the following information carefully and discuss it with others if you wish to and ask questions about any of the information before you decide to participate. You may also wish to talk to your family or friends about this study, before agreeing to join.

**Key Information for You to Consider**

- **Voluntary Consent**. You are being asked to volunteer for a research study. You can choose whether you would like to participate or not. If you do agree you can change your mind at any time and withdraw from the research. This will not affect your work now or in the future.
- **Purpose**. We are doing this research to better understand how health workers and teams work together to deliver neonatal care, including how changes might impact on different people in the team and how communication might be improved. The research will be used to inform how quality improvement interventions are implemented in hospitals. You have been invited to participate in this research because you work in neonatal care, or you work in the hospital and support neonatal care in some way (the delivery unit, post-natal unit, neonatal unit and paediatric unit, laboratory, pharmacy, nutrition, patient support, physiotherapy, orthopaedic/plaster care and occupational therapy).
- **Duration.** Your participation in this study will last 45-60 minutes.
- **Procedures and Activities.** We will ask you some questions for you to respond to. These discussions will either be online, via phone or face-to-face. If conducted face-to-face, the requisite Ministry of Health COVID-19 protocols will be adhered to. If you agree to participate the researcher will ask you to sign a consent form before asking any questions. You can stop or pause the in-depth interview at any time, by letting the researcher know.

**Who is carrying out this study and what is this study about?**

- This study is being carried out by KEMRI in collaboration with the University of Oxford, UK. KEMRI is a Kenyan government organization that carries out medical research to find better ways of preventing and treating illness in the future for everybody’s benefit.
- The study involves asking health workers what they know, feel or do when they need help or advice from colleagues, using existing social interactions networks and how these networks and social interactions impact care of neonates.
- We want to better understand how health workers and teams work together to deliver neonatal care, including how changes might impact on different people in the team and how communication might be improved.

**Why do you want to talk to me and what does it involve?**

- *Selection*

This hospital has several clinical environments where neonates receive care, including many supportive departments that contribute to the care of neonates. You have been selected from your department/unit so that we get experiences from individuals that will give us a true picture of the social networks and connections in the hospital, without preference.

- *About the In-depth interview*:

I/my colleague would like to ask you a number of questions about the nature of social connections and interactions between and among health workers, how they are formed and why and how this impacts care of neonates.

The interview will be audio-recorded (audio or video recorded if using an online platform such as MS Teams) to assist later in fully writing up the information. A researcher may sometimes need to contact you again, to clarify information.

**Are there any risks or disadvantages to me for taking part?**

- There are no risks for taking part in the study. However, questions around interactions and social connections can be sensitive as they may include a discussion about difficulty relationships at workplace.
- We will aim to conduct the interview during your tea or lunch break or when you are free while you are on duty to minimize disruption of your duties and obligation to work.
- If you are attending this in-depth interview while on your off or leave day (and not on a working day/shift), we will reimburse your transport costs at a rate of Ksh. 600 per person or mileage costs at the rate of Ksh.25/km for the face-to-face interview
- Online interviews will be reimbursed with a data voucher worth Ksh.250 while phone interviews will be at the cost of the researcher.

**Are there any advantages to me for taking part?**

- There are no individual benefits to taking part in this study. In participating in this study, you will contribute to knowledge of social connections and ties, including knowledge of factors that facilitate or hinder use of knowledge and skills acquired during nurse trainings. This finding may help in making recommendations in the design of interventions that improve quality of care of neonates in Kenya and elsewhere in the future, for example through developing new health policies.

**Data Protection**

**Who will have access to information about me in this research?**

- All our research records are stored securely in locked cabinets and password protected computers and are accessed only by authorized persons.
- The personally identifiable information you provide will be used only for study purposes such as contact you for scheduling of visits and verification of consent through signed consent forms.
- We will share anonymized individual and summary information we collect or generate with our partner institution, University of Oxford in ways that do not reveal individual participants’ identities.
- Anonymised data will be stored in a secure KEMRI-Wellcome Trust Research Programme repository and may be used for future research.
- KEMRI will keep any keep any personally identifiable information about you from this study for 10 years after the study has finished in accordance with applicable Data Protection Requirements both in Kenya and in the UK. You have the right to access the personal data we hold that pertains to you, to object to or make corrections to the processing of all or part of the personal data. However, this might be limited due to coding which might make unblinding difficult.

**Will the research be published? Could I be identified from any publications or other research outputs?**

- The research will be published. We might however include direct quotations, but without identifying you, in any research outputs.
- In future, information collected or generated during this study may be used to support new research by other researchers in Kenya or other countries on improving newborn service delivery. In all cases, we will only share information with other researchers in ways that do not reveal individual participants’ identities. For example, we will remove such as their names and where they live and replace this information with number codes. Any future research using information from this study must first be approved by a local or national expert committee to make sure that the interests of participants and their communities are protected.
- University of Oxford is responsible for ensuring that staff involved in the study in Kenya adhere to the safe and proper use of any personal information you provide. You can contact the research team for any further information about how your data will be managed. Further information about your rights with respect to your personal data is available from <https://compliance.admin.ox.c.uk/individual-rights>

**Who has allowed this research to take place?**

All research at KEMRI must be approved before it begins by an independent national committee, an international committee and a committee at the KEMRI Wellcome Trust Research Programme. The committees look carefully at planned work before it begins, and must agree that the research is important, relevant to Kenya and follows nationally and internationally agreed research guidelines. This includes ensuring that all participants’ safety and rights are respected. This study will also seek ethics approval from a subcommittee of the University of Oxford Central University Research Ethics Committee.

**What will happen if I refuse to participate?**

All participation in research is voluntary. You are free to decide if you want to take part or not. If you do agree you can change your mind at any time without any consequences. You can withdraw yourself from the study, without giving a reason, and without negative consequences, by advising us of this decision. The deadline by which you can withdraw from the study is September 2022. because we expect that data from this study would have already began being analysed by this time. If you withdraw from the study after this date, all information that you have already provided will still be included in the analysis. If you withdraw from the study before this date, information you have already provided may still be included if analysis has already occurred, however the information you have provided will not be used as quotes and the information will be removed from any data repositories.

**What if I have any questions?**

You are free to ask me any question about this research. If you have any further questions about the study, you are free to contact the research team using the contacts below:

Mr. Conrad Wanyama, KEMRI Wellcome Trust Research Programme, P.O. Box 230, Kilifi. Telephone: [mobile 0721275262] or 0700274694

**If you want to ask someone independent anything about this research please contact:**

Community Liaison Manager, KEMRI Wellcome Trust Research Programme, P.O. Box 230, Kilifi. Telephone: 041 7522 063, Mobile 0723 342 780 or 0705 154 386

***And***

### KEMRI-Wellcome Trust Research Programme consent form for In-depth interview for health workers for: -

**Study title: Pathways Study: Understanding health worker social ties and communication, and their influence in neonatal care.**

**Lay Title: Understanding how health worker interactions influence the care of newborn babies.**

I have had the study explained to me. I have understood all that has been read/explained and had my questions answered satisfactorily. And I agree to take part in this research

**Please initial the sentences that reflect your choices, and then sign below:**

I do wish to be notified by investigators in the event of research findings of possible importance to my institution or myself. **Yes	No**

I agree that the study team use the identifier that I have provided (telephone number, country ID number, etc.) to locate me in the future. **Yes	No**

I agree for the interview to be recorded **Yes	No**

I agree to the use of quotations in research outputs if I am not identifiable **Yes	No**

I agree to my anonymised research data being used, for future research **Yes	No**

I understand that I can change my mind at any stage.

| **Signature:** |  | **Date:** |
| --- | --- | --- |
| **Participant Name:** |  | **Time:** |
|  | (Please print name) |  |

--------------------------------------------------------------------------------------------------------------------------------------

I have followed the study procedure to obtain consent from the participant. S/he apparently understood the nature and the purpose of the study and consents to the participation in the study. S/he has been given opportunity to ask questions which have been answered satisfactorily.

**Designee/investigator’s signature:** ____________________________ **Date** ____________

**Designee/investigator’s name:** _____________________________**Time** ____________

(Please print name)

*THE PARTICIPANT SHOULD NOW BE GIVEN A SIGNED COPY TO KEEP*

*……………………………………………………………………………………………………………………………………………………………………………*

### Appendix M: KEMRI-Wellcome Trust Research Programme: Participant Information Sheet and Consent form for key informant interviews for: -

**Pathways Study Form, M: Key informant interview (for nursing managers and unit leaders in neonatal care units or related environments)**

**Study title: Pathways Study: Understanding health worker social ties and communication, and their influence in neonatal care.**

**Lay Title: Understanding how health worker interactions influence the care of newborn babies.**

| Institution | Investigators |
| --- | --- |
| KEMRI Wellcome Trust Research Programme | Conrad Wanyama, Prof. Mike English, Dr. Jalemba Aluvaala, Prof Grace Irimu, Joyline Jepkosgei, Peris Musitia, Dr. Dorothy Oluoch, Prof Sassy Molyneux |
| Nuffield Department of Medicine, University of Oxford, UK | Dr Claire Blacklock Dr. Jacob McKnight*,* |
| THIS Institute, University of Cambridge | Prof. Lisa Hinton |
| Nuffield Department of Primary Care Health Sciences, University of Oxford, UK | Dr. Geoff Wong |
| University of Nairobi | Dr. Aluvaala Jalemba, Dr. Joyce Jebet, Prof Grace Irimu |
| Kenyatta National Hospital | Catherine Ngugi, Josephine Bariu |
| Mbagathi County Referral Hospital | Norah Chebet |
| Kenya Medical Training College | Everlyne Abuga |

You are being asked to take part in a study. The box below tells you important things you should think about before deciding to join the study. We will provide more detailed information below the box. Please take time to read the following information carefully and discuss it with others if you wish to and ask questions about any of the information before you decide to participate. You may also wish to talk to your family or friends about this study, before agreeing to join.

**Key Information for You to Consider**

- **Voluntary Consent**. You are being asked to volunteer for a research study. You can choose whether you would like to participate or not. If you do agree you can change your mind at any time and withdraw from the research. This will not affect your work now or in the future.
- **Purpose**. We are doing this research to better understand how health workers and teams work together to deliver neonatal care, including how changes might impact on different people in the team and how communication might be improved. The research will be used to inform how quality improvement interventions are implemented in hospitals. You have been invited to participate in this research because you are one of the nurse managers or unit leaders with experience managing the facility and providing leadership to junior colleagues (nurses and nurse-students).
- **Duration.** Your participation in this study will last 30-45 minutes.
- **Procedures and Activities.** We will ask you some questions for you to respond to while in a quiet, private place. These discussions will either be online, via phone or face-to-face. If conducted face-to-face, the requisite Ministry of Health COVID-19 protocols will be adhered to. If you agree to participate the researcher will ask you to sign a consent form before asking any questions. You can stop or pause the key informant interview at any time, by letting the researcher know. We will audio-record your answers using a Dictaphone or online software in line with organisational policies (e.g. MS Teams). If conducted online or via phone, the conversation will be audio-recorded on the organizational device (online software such as MS Teams or institutional smartphone).

**Who is carrying out this study and what is this study about?**

- This study is being carried out by KEMRI in collaboration with the University of Oxford, UK. KEMRI is a Kenyan government organization that carries out medical research to find better ways of preventing and treating illness in the future for everybody’s benefit.
- The study involves asking you information about your unit, how nursing care is planned and provided to newborn babies and nursing staff view the role of non-technical/socio-technical skills, including social interactions and networks in the care of neonates.

**Why do you want to talk to me and what does it involve?**

- *Selection*

You have been selected to participate for this interview because we feel your current position and experience can contribute much to our understanding and knowledge of the non-technical (human) factors that either hinder or facilitate how nurses use the clinical knowledge and skills they have acquired during training in the care of neonates at the workplace.

- *About the key informant interview*:

I/my colleague would like to ask you a number of questions about the work environment and how care is planned for neonates that is unique to your care context.

The interview will be audio-recorded (audio or video recorded if using an online platform such as MS Teams) to assist later in fully writing up the information. A researcher may sometimes need to contact you again, to clarify information.

**Are there any risks or disadvantages to me for taking part?**

- There are no risks for taking part in the study. We will aim not to link this information with your unit or hospital during future work (analysis and publications)
- We will aim to conduct the interview during your tea or lunch break or when you are free while you are on duty to minimize disruption of your duties and obligation to work.
- If you are attending this key informant interview while on your off or leave day (and not on a working day/shift), we will reimburse your transport costs at a rate of Ksh. 600 per person or mileage costs at the rate of Ksh.25/km for the face-to-face interview
- Online interviews will be reimbursed with a data voucher worth Ksh.250 while phone interviews will be at the cost of the researcher.

**Are there any advantages to me for taking part?**

- There are no individual benefits to taking part in this study. In participating in this study, you will contribute to knowledge of social connections and ties, including knowledge of factors that facilitate or hinder use of knowledge and skills acquired during nurse trainings. This finding may help in making recommendations in the design of interventions that improve quality of care of neonates in Kenya and elsewhere in the future, for example through developing new health policies.

**Data Protection**

**Who will have access to information about me in this research?**

- All our research records are stored securely in locked cabinets and password protected computers and are accessed only by authorized persons.
- The personally identifiable information you provide will be used only for study purposes such as contact you for scheduling of visits and verification of consent through signed consent forms.
- We will share anonymized individual and summary information we collect or generate with our partner institution, University of Oxford in ways that do not reveal individual participants’ identities.
- Anonymised data will be stored in a secure KEMRI-Wellcome Trust Research Programme repository and may be used for future research.
- KEMRI will keep any keep any personally identifiable information about you from this study for 10 years after the study has finished in accordance with applicable Data Protection Requirements both in Kenya and in the UK. You have the right to access the personal data we hold that pertains to you, to object to or make corrections to the processing of all or part of the personal data. However, this might be limited due to coding which might make unblinding difficult.

**Will the research be published? Could I be identified from any publications or other research outputs?**

- The research will be published. We might however include direct quotations, but without identifying you, in any research outputs.
- In future, information collected or generated during this study may be used to support new research by other researchers in Kenya or other countries on improving newborn service delivery. In all cases, we will only share information with other researchers in ways that do not reveal individual participants’ identities. For example, we will remove such as their names and where they live and replace this information with number codes. Any future research using information from this study must first be approved by a local or national expert committee to make sure that the interests of participants and their communities are protected.
- University of Oxford is responsible for ensuring that staff involved in the study in Kenya adhere to the safe and proper use of any personal information you provide. You can contact the research team for any further information about how your data will be managed. Further information about your rights with respect to your personal data is available from <https://compliance.admin.ox.c.uk/individual-rights>

**Who has allowed this research to take place?**

All research at KEMRI must be approved before it begins by an independent national committee, an international committee and a committee at the KEMRI Wellcome Trust Research Programme. The committees look carefully at planned work before it begins, and must agree that the research is important, relevant to Kenya and follows nationally and internationally agreed research guidelines. This includes ensuring that all participants’ safety and rights are respected. This study will also seek ethics approval from a subcommittee of the University of Oxford Central University Research Ethics Committee.

**What will happen if I refuse to participate?**

All participation in research is voluntary. You are free to decide if you want to take part or not. If you do agree you can change your mind at any time without any consequences. You can withdraw yourself from the study, without giving a reason, and without negative consequences, by advising us of this decision. The deadline by which you can withdraw from the study is September 2022, because we expect that data from this study would have already began being analysed by this time. If you withdraw from the study after this date, all information that you have already provided will still be included in the analysis. If you withdraw from the study before this date, information you have already provided may still be included if analysis has already occurred, however the information you have provided will not be used as quotes and the information will be removed from any data repositories.

**What if I have any questions?**

You are free to ask me any question about this research. If you have any further questions about the study, you are free to contact the research team using the contacts below:

Mr. Conrad Wanyama, KEMRI Wellcome Trust Research Programme, P.O. Box 230, Kilifi. Telephone: [mobile 0721275262] or 0700274694

**If you want to ask someone independent anything about this research please contact:**

Community Liaison Manager, KEMRI Wellcome Trust Research Programme, P.O. Box 230, Kilifi. Telephone: 041 7522 063, Mobile 0723 342 780 or 0705 154 386

***And***

### KEMRI-Wellcome Trust Research Programme consent form for key informant interview (hospital nurse managers) for: -

**Study title: Pathways Study: Understanding health worker social ties and communication, and their influence in neonatal care.**

**Lay Title: Understanding how health worker interactions influence the care of newborn babies.**

I have had the study explained to me. I have understood all that has been read/explained and had my questions answered satisfactorily. And I agree to take part in this research

**Please initial the sentences that reflect your choices, and then sign below:**

I do wish to be notified by investigators in the event of research findings of possible importance to my institution or myself. **Yes	No**

I agree that the study team use the identifier that I have provided (telephone number, country ID number, etc.) to locate me in the future. **Yes	No**

I agree for the interview to be recorded **Yes	No**

I agree to the use of quotations in research outputs if I am not identifiable **Yes	No**

I agree to my anonymised research data being used, for future research **Yes	No**

I understand that I can change my mind at any stage.

| **Signature:** |  | **Date:** |
| --- | --- | --- |
| **Participant Name:** |  | **Time:** |
|  | (Please print name) |  |

--------------------------------------------------------------------------------------------------------------------------------------

I have followed the study procedure to obtain consent from the participant. S/he apparently understood the nature and the purpose of the study and consents to the participation in the study. S/he has been given opportunity to ask questions which have been answered satisfactorily.

**Designee/investigator’s signature:** ____________________________ **Date** ____________

**Designee/investigator’s name:** _____________________________**Time** ____________

(Please print name)

*THE PARTICIPANT SHOULD NOW BE GIVEN A SIGNED COPY TO KEEP*

*……………………………………………………………………………………………………………………………………………………………………………*

### Appendix N– KEMRI-Wellcome Trust Research Programme: Participant Information Sheet and Consent form for key informant interviews for: -

**Pathways Study Form N: Key informant interview (for nursing educators in training institutions)**

**Study title: Pathways Study: Understanding health worker social ties and communication, and their influence in neonatal care.**

**Lay Title: Understanding how health worker interactions influence the care of newborn babies.**

| Institution | Investigators |
| --- | --- |
| KEMRI Wellcome Trust Research Programme | Conrad Wanyama, Prof. Mike English, Dr. Jalemba Aluvaala, Prof Grace Irimu, Joyline Jepkosgei, Peris Musitia, Dr. Dorothy Oluoch, Prof Sassy Molyneux |
| Nuffield Department of Medicine, University of Oxford, UK | Dr Claire Blacklock Dr. Jacob McKnight*,* |
| THIS Institute, University of Cambridge | Prof. Lisa Hinton |
| Nuffield Department of Primary Care Health Sciences, University of Oxford, UK | Dr. Geoff Wong |
| University of Nairobi | Dr. Aluvaala Jalemba, Dr. Joyce Jebet, Prof Grace Irimu |
| Kenyatta National Hospital | Catherine Ngugi, Josephine Bariu |
| Mbagathi County Referral Hospital | Norah Chebet |
| Kenya Medical Training College | Everlyne Abuga |

You are being asked to take part in a study. The box below tells you important things you should think about before deciding to join the study. We will provide more detailed information below the box. Please take time to read the following information carefully and discuss it with others if you wish to and ask questions about any of the information before you decide to participate. You may also wish to talk to your family or friends about this study, before agreeing to join.

**Key Information for You to Consider**

- **Voluntary Consent**. You are being asked to volunteer for a research study. You can choose whether you would like to participate or not. If you do agree you can change your mind at any time and withdraw from the research. This will not affect your work now or in the future.
- **Purpose**. We are doing this research to better understand how socio-cognitive skills influence the use of nurses’ clinical competencies in neonatal care in Kenya as well as how health workers and teams work together to deliver neonatal care, including how changes might impact on different people in the team. The research will be used to inform how education-specific quality improvement interventions are designed and implemented in training institutes and hospitals. You have been invited to participate in this research because you are one of the nurse educators with leadership and/or programme management roles with experience in designing and implementing trainings to nursing students.
- **Duration.** Your participation in this study will last 30-45 minutes.
- **Procedures and Activities.** We will ask you some questions for you to respond to while in a quiet, private place. These discussions will either be online, via phone or face-to-face. If conducted face-to-face, the requisite Ministry of Health COVID-19 protocols will be adhered to. If you agree to participate the researcher will ask you to sign a consent form before asking any questions. You can stop or pause the key informant interview at any time, by letting the researcher know. We will audio-record your answers using a Dictaphone or online software in line with organisational policies (e.g. MS Teams). If conducted online or via phone, the conversation will be audio-recorded on the organizational device (online software such as MS Teams or institutional smartphone).

**Who is carrying out this study and what is this study about?**

- This study is being carried out by KEMRI in collaboration with the University of Oxford, UK. KEMRI is a Kenyan government organization that carries out medical research to find better ways of preventing and treating illness in the future for everybody’s benefit.
- The study involves asking you information about your unit, how nursing care is planned and provided to newborn babies and nursing staff view the role of non-technical/socio-technical skills, including social interactions snd networks in the care of neonates.

**Why do you want to talk to me and what does it involve?**

- *Selection*

You have been selected to participate for this interview because we feel your current position and experience can contribute much to our understanding and knowledge of the non-technical (human) factors that either hinder or facilitate how nurses use the knowledge and skills they have acquired during training at the workplace.

- *About the key informant interview*:

I/my colleague would like to ask you a number of questions about how training for nurses is designed and implemented around neonatal care and specific to your training institutional context.

The interview will be audio-recorded (audio or video recorded if using an online platform such as MS Teams) to assist later in fully writing up the information. A researcher may sometimes need to contact you again, to clarify information.

**Are there any risks or disadvantages to me for taking part?**

- There are no risks for taking part in the study. Information collected during this interview will not be linked to you or your institution during analysis and publication.
- We will aim to conduct the interview during your tea or lunch break or when you are free while you are on duty to minimize disruption of your duties and obligation to work. If we conduct these during tea/lunch break, we will provide refreshments.
- If you are attending this key informant interview while on your off or leave day (and not on a working day/shift), we will reimburse your transport costs at a rate of Ksh. 600 per person or mileage costs at the rate of Ksh.25/km for the face-to-face interview
- Online questionnaire interviews will be reimbursed with a data voucher worth Ksh.250-500. Phone interviews will be at the cost of the researcher.

**Are there any advantages to me for taking part?**

- There are no individual benefits to taking part in this study. In participating in this study, you will contribute to the influence of socio-cognitive factors as either facilitators or hinderances to the use of knowledge and skills acquired during nurse trainings. This finding may help in making recommendations in the design of interventions that improve quality of care of neonates in Kenya and elsewhere in the future, for example through developing new health policies.

**Data Protection**

**Who will have access to information about me in this research?**

- All our research records are stored securely in locked cabinets and password protected computers and are accessed only by authorized persons.
- The personally identifiable information you provide will be used only for study purposes such as contact you for scheduling of visits and verification of consent through signed consent forms.
- We will share anonymized individual and summary information we collect or generate with our partner institution, University of Oxford in ways that do not reveal individual participants’ identities.
- Anonymised data will be stored in a secure KEMRI-Wellcome Trust Research Programme repository and may be used for future research.
- KEMRI will keep any keep any personally identifiable information about you from this study for 10 years after the study has finished in accordance with applicable Data Protection Requirements both in Kenya and in the UK. You have the right to access the personal data we hold that pertains to you, to object to or make corrections to the processing of all or part of the personal data. However, this might be limited due to coding which might make unblinding difficult.

**Will the research be published? Could I be identified from any publications or other research outputs?**

- The research will be published. We might however include direct quotations, but without identifying you, in any research outputs.
- In future, information collected or generated during this study may be used to support new research by other researchers in Kenya or other countries on improving newborn service delivery. In all cases, we will only share information with other researchers in ways that do not reveal individual participants’ identities. For example, we will remove such as their names and where they live and replace this information with number codes. Any future research using information from this study must first be approved by a local or national expert committee to make sure that the interests of participants and their communities are protected.
- University of Oxford is responsible for ensuring that staff involved in the study in Kenya adhere to the safe and proper use of any personal information you provide. You can contact the research team for any further information about how your data will be managed. Further information about your rights with respect to your personal data is available from <https://compliance.admin.ox.c.uk/individual-rights>

**Who has allowed this research to take place?**

All research at KEMRI must be approved before it begins by an independent national committee, an international committee and a committee at the KEMRI Wellcome Trust Research Programme. The committees look carefully at planned work before it begins, and must agree that the research is important, relevant to Kenya and follows nationally and internationally agreed research guidelines. This includes ensuring that all participants’ safety and rights are respected. This study will also seek ethics approval from a subcommittee of the University of Oxford Central University Research Ethics Committee.

**What will happen if I refuse to participate?**

All participation in research is voluntary. You are free to decide if you want to take part or not. If you do agree you can change your mind at any time without any consequences. You can withdraw yourself from the study, without giving a reason, and without negative consequences, by advising us of this decision. The deadline by which you can withdraw from the study is September 2022, because we expect that data from this study would have already began being analysed by this time. If you withdraw from the study after this date, all information that you have already provided will still be included in the analysis. If you withdraw from the study before this date, information you have already provided may still be included if analysis has already occurred, however the information you have provided will not be used as quotes and the information will be removed from any data repositories.

**What if I have any questions?**

You are free to ask me any question about this research. If you have any further questions about the study, you are free to contact the research team using the contacts below:

Mr. Conrad Wanyama, KEMRI Wellcome Trust Research Programme, P.O. Box 230, Kilifi. Telephone: [mobile 0721275262] or 0700274694

**If you want to ask someone independent anything about this research please contact:**

Community Liaison Manager, KEMRI Wellcome Trust Research Programme, P.O. Box 230, Kilifi. Telephone: 041 7522 063, Mobile 0723 342 780 or 0705 154 386

***And***

### KEMRI-Wellcome Trust Research Programme consent form for key informant interview (College and School Managers/educators) for: -

**Study title: Pathways Study: Understanding Health Worker Communication Pathways and their influence in Neonatal Care**

**Lay Title: Understanding how health worker interactions influence the care of newborn babies.**

I have had the study explained to me. I have understood all that has been read/explained and had my questions answered satisfactorily. And I agree to take part in this research

**Please initial the sentences that reflect your choices, and then sign below:**

I do wish to be notified by investigators in the event of research findings of possible importance to my institution or myself. **Yes	No**

I agree that the study team use the identifier that I have provided (telephone number, country ID number, etc.) to locate me in the future. **Yes	No**

I agree for the interview to be recorded	**Yes	No**

I agree to the use of quotations in research outputs if I am not identifiable **Yes	No**

I agree to my anonymised research data being used, for future research **Yes	No**

I understand that I can change my mind at any stage.

| **Signature:** |  | **Date:** |
| --- | --- | --- |
| **Participant Name:** |  | **Time:** |
|  | (Please print name) |  |

--------------------------------------------------------------------------------------------------------------------------------------

I have followed the study procedure to obtain consent from the participant. S/he apparently understood the nature and the purpose of the study and consents to the participation in the study. S/he has been given opportunity to ask questions which have been answered satisfactorily.

**Designee/investigator’s signature:** ____________________________ **Date** ____________

**Designee/investigator’s name:** _____________________________**Time** ____________

(Please print name)

*THE PARTICIPANT SHOULD NOW BE GIVEN A SIGNED COPY TO KEEP*

*……………………………………………………………………………………………………………………………………………………………………………*

### Appendix O– KEMRI-Wellcome Trust Research Programme: Participant Information Sheet and Consent form for Focus Group Discussion for: -

**Pathways Study Form O: Focus Group Discussion (for nursing graduates and nurse educators)**

**Study title: Pathways Study: Understanding health worker social ties and communication, and their influence in neonatal care.**

**Lay Title: Understanding how health worker interactions influence the care of newborn babies.**

| Institution | Investigators |
| --- | --- |
| KEMRI Wellcome Trust Research Programme | Conrad Wanyama, Prof. Mike English, Dr. Jalemba Aluvaala, Prof Grace Irimu, Joyline Jepkosgei, Peris Musitia, Dr. Dorothy Oluoch, Prof Sassy Molyneux |
| Nuffield Department of Medicine, University of Oxford, UK | Dr Claire Blacklock Dr. Jacob McKnight*,* |
| THIS Institute, University of Cambridge | Prof. Lisa Hinton |
| Nuffield Department of Primary Care Health Sciences, University of Oxford, UK | Dr. Geoff Wong |
| University of Nairobi | Dr. Aluvaala Jalemba, Dr. Joyce Jebet, Prof Grace Irimu |
| Kenyatta National Hospital | Catherine Ngugi, Josephine Bariu |
| Mbagathi County Referral Hospital | Norah Chebet |
| Kenya Medical Training College | Everlyne Abuga |

You are being asked to take part in a study. The box below tells you important things you should think about before deciding to join the study. We will provide more detailed information below the box. Please take time to read the following information carefully and discuss it with others if you wish to and ask questions about any of the information before you decide to participate. You may also wish to talk to your family or friends about this study, before agreeing to join.

**Key Information for You to Consider**

- **Voluntary Consent**. You are being asked to volunteer for a research study. You can choose whether you would like to participate or not. If you do agree you can change your mind at any time and withdraw from the research. This will not affect your work now or in the future.
- **Purpose**. We are doing this research to better understand human and other workplace factors (also called non-technical or socio-cognitive skills) that either facilitate or hinder the use of skills and knowledge acquired during nurse training. The research will be used to inform how education-specific quality improvement interventions are designed and implemented in training institutes and hospitals. You have been invited to participate in this research because you are one of the nurse graduates providing clinical care for neonates in Kenya.
- **Duration.** Your participation in this study will last 60-90 minutes.
- **Procedures and Activities.** We will ask you some questions, in a group of 5-9 members, for you to respond to while in a quiet, private place. These discussions will either be online or face-to-face. If conducted face-to-face, the requisite Ministry of Health COVID-19 protocols will be adhered to. If you agree to participate the researcher will ask you to sign a consent form before asking any questions. You can stop or pause the discussion at any time, by letting the researcher know. We will audio-record your answers using a Dictaphone or online software in line with organisational policies (e.g. MS Teams). If conducted online the meeting conversations will be audio-recorded on the organizational device (using online software such as MS Teams).

**Who is carrying out this study and what is this study about?**

- This study is being carried out by KEMRI in collaboration with the University of Oxford, UK. KEMRI is a Kenyan government organization that carries out medical research to find better ways of preventing and treating illness in the future for everybody’s benefit.
- The study involves asking you information about human and other workplace factors (non-technical/socio-technical skills) that either facilitate or hinder the use of skills and knowledge acquired during training.

**Why do you want to talk to me and what does it involve?**

- *Selection*

You have been selected to participate in this focus group discussion because we fill you can provide relevant information based on your experience as well as clinical cadre while previously as a nurse student and now as a nurse graduate providing clinical care to neonates in Kenya. We are interested in knowing what you know and about availability and how non-technical human and workplace factors (socio-cognitive skills) either hinder or facilitate how nurses use their clinical knowledge and skills they have acquired during training at the workplace.

- *About the focus group discussion*:

I/my colleague would like to ask you, within the group, a number of questions about the influence of socio-cognitive skills in the use of clinical knowledge and skills among nurses providing care to neonates.

The discussion will be audio-recorded (audio or video recorded if using an online platform such as MS Teams) to assist later in fully writing up the information. A researcher may sometimes need to contact you again, to clarify information.

**Are there any risks or disadvantages to me for taking part?**

- There are no risks for taking part in the study. Information collected during this discussion will not be linked to you or your institution during analysis and publication.
- We will aim to conduct the discussion during your tea or lunch break or when you are free while you are on duty to minimize disruption of your duties and obligation to work.
- If you are attending this focus group discussion while on your off or leave day (and not on a working day/shift), we will reimburse your transport costs at a rate of Ksh. 600 per person or mileage costs at the rate of Ksh.25/km for the face-to-face FGD meeting.
- Online FGD meetings will be reimbursed with a data voucher worth Ksh.250-500.

**Are there any advantages to me for taking part?**

- There are no individual benefits to taking part in this study. In participating in this study, you will contribute to the influence of socio-cognitive factors as either facilitators or hinderances to the use of knowledge and skills acquired during nurse trainings. This finding may help in making recommendations in the design of interventions that improve quality of care of neonates in Kenya and elsewhere in the future, for example through developing new health policies.

**Data Protection**

**Who will have access to information about me in this research?**

- All our research records are stored securely in locked cabinets and password protected computers and are accessed only by authorized persons.
- The personally identifiable information you provide will be used only for study purposes such as contact you for scheduling of visits and verification of consent through signed consent forms.
- We will share anonymized individual and summary information we collect or generate with our partner institution, University of Oxford in ways that do not reveal individual participants’ identities.
- Anonymised data will be stored in a secure KEMRI-Wellcome Trust Research Programme repository and may be used for future research.
- KEMRI will keep any keep any personally identifiable information about you from this study for 10 years after the study has finished in accordance with applicable Data Protection Requirements both in Kenya and in the UK. You have the right to access the personal data we hold that pertains to you, to object to or make corrections to the processing of all or part of the personal data. However, this might be limited due to coding which might make unblinding difficult.

**Will the research be published? Could I be identified from any publications or other research outputs?**

- The research will be published. We might however include direct quotations, but without identifying you, in any research outputs.
- In future, information collected or generated during this study may be used to support new research by other researchers in Kenya or other countries on improving newborn service delivery. In all cases, we will only share information with other researchers in ways that do not reveal individual participants’ identities. For example, we will remove such as their names and where they live and replace this information with number codes. Any future research using information from this study must first be approved by a local or national expert committee to make sure that the interests of participants and their communities are protected.
- University of Oxford is responsible for ensuring that staff involved in the study in Kenya adhere to the safe and proper use of any personal information you provide. You can contact the research team for any further information about how your data will be managed. Further information about your rights with respect to your personal data is available from <https://compliance.admin.ox.c.uk/individual-rights>

**Who has allowed this research to take place?**

All research at KEMRI must be approved before it begins by an independent national committee, an international committee and a committee at the KEMRI Wellcome Trust Research Programme. The committees look carefully at planned work before it begins, and must agree that the research is important, relevant to Kenya and follows nationally and internationally agreed research guidelines. This includes ensuring that all participants’ safety and rights are respected. This study will also seek ethics approval from a subcommittee of the University of Oxford Central University Research Ethics Committee.

**What will happen if I refuse to participate?**

All participation in research is voluntary. You are free to decide if you want to take part or not. If you do agree you can change your mind at any time without any consequences. You can withdraw yourself from the study, without giving a reason, and without negative consequences, by advising us of this decision. The deadline by which you can withdraw from the study is September 2022, because we expect that data from this study would have already began being analysed by this time. If you withdraw from the study after this date, all information that you have already provided will still be included in the analysis. If you withdraw from the study before this date, information you have already provided may still be included if analysis has already occurred, however the information you have provided will not be used as quotes and the information will be removed from any data repositories.

**What if I have any questions?**

You are free to ask me any question about this research. If you have any further questions about the study, you are free to contact the research team using the contacts below:

Mr. Conrad Wanyama, KEMRI Wellcome Trust Research Programme, P.O. Box 230, Kilifi. Telephone: [mobile 0721275262] or 0700274694

**If you want to ask someone independent anything about this research please contact:**

Community Liaison Manager, KEMRI Wellcome Trust Research Programme, P.O. Box 230, Kilifi. Telephone: 041 7522 063, Mobile 0723 342 780 or 0705 154 386

***And***

### KEMRI-Wellcome Trust Research Programme consent form for Focus group Discussion (nurse graduates and nurse educators) for: -

**Study title: Pathways Study: Understanding health worker social ties and communication, and their influence in neonatal care.**

**Lay Title: Understanding how health worker interactions influence the care of newborn babies.**

I have had the study explained to me. I have understood all that has been read/explained and had my questions answered satisfactorily. And I agree to take part in this research

**Please initial the sentences that reflect your choices, and then sign below:**

I do wish to be notified by investigators in the event of research findings of possible importance to my professional group or myself. **Yes	No**

I agree that the study team use the identifier that I have provided (telephone number, country ID number, etc.) to locate me in the future. **Yes	No**

**I agree for the interview to be recorded Yes	No**

I agree to the use of quotations in research outputs if I am not identifiable **Yes	No**

I agree to my anonymised research data being used, for future research **Yes	No**

I understand that I can change my mind at any stage.

| **Signature:** |  | **Date:** |
| --- | --- | --- |
| **Participant Name:** |  | **Time:** |
|  | (Please print name) |  |

--------------------------------------------------------------------------------------------------------------------------------------

I have followed the study procedure to obtain consent from the participant. S/he apparently understood the nature and the purpose of the study and consents to the participation in the study. S/he has been given opportunity to ask questions which have been answered satisfactorily.

**Designee/investigator’s signature:** ____________________________ **Date** ____________

**Designee/investigator’s name:** _____________________________**Time** ____________

(Please print name)

*THE PARTICIPANT SHOULD NOW BE GIVEN A SIGNED COPY TO KEEP*

*……………………………………………………………………………………………………………………………………………………………………………*

### Appendix P – KEMRI-Wellcome Trust Research Programme: Participant Information Sheet and verbal assent form for stakeholder workshop for:

**Pathways Study Form P: Stakeholder workshop**

**Study title: Pathways Study: Understanding health worker social ties and communication, and their influence in neonatal care.**

**Lay Title: Understanding how health worker interactions influence the care of newborn babies.**

| Institution | Investigators |
| --- | --- |
| KEMRI Wellcome Trust Research Programme | Conrad Wanyama, Prof. Mike English, Dr. Jalemba Aluvaala, Prof Grace Irimu, Joyline Jepkosgei, Peris Musitia, Dr. Dorothy Oluoch, Prof Sassy Molyneux |
| Nuffield Department of Medicine, University of Oxford, UK | Dr Claire Blacklock Dr. Jacob McKnight*,* |
| THIS Institute, University of Cambridge | Prof. Lisa Hinton |
| Nuffield Department of Primary Care Health Sciences, University of Oxford, UK | Dr. Geoff Wong |
| University of Nairobi | Dr. Aluvaala Jalemba, Dr. Joyce Jebet, Prof Grace Irimu |
| Kenyatta National Hospital | Catherine Ngugi, Josephine Bariu |
| Mbagathi County Referral Hospital | Norah Chebet |
| Kenya Medical Training College | Everlyne Abuga |

You are being invited to take part in a research project. Before you decide it is important for you to understand why the research is being done and what it will involve. Please take time to read the following information and ask us if there is anything that is not clear or if you would like more information. Take time to decide whether you wish to take part.

**Key Information for You to Consider**

- **Voluntary Consent**. You are being asked to volunteer for a research study. You can choose whether you would like to participate or not. If you do agree you can change your mind at any time and withdraw from the research. This will not affect your work now or in the future.
- **Purpose**. We are doing this research to better understand how health workers and teams work together to deliver neonatal care, including how changes might impact on different people in the team and how communication might be improved. We also are interested in understanding the role of non-technical factors (socio-cognitive skills) in facilitating or hindering the use of clinical competencies among nurses working in neonatal areas. The research will be used to inform how quality improvement interventions are implemented in hospitals and training institutions. We have invited you because you have relevant experience in healthcare delivery (particularly neonatal care), research and policy/decision making, and you are familiar with key aspects of intervention design or implementation.
- **Duration.** Your participation in this event will last 5-7 hours.
- **Procedures and Activities.** You will participate in this one-day stakeholder workshop event to contribute to group discussions with other participants and the research team, to make sense of and comment and reflect on the data. You will also help in the co-design of a set of recommendations, as an output from the workshop. This event will be audio-recorded. If conducted online the meeting conversations will be audio-recorded on the organizational device (using online software such as MS Teams).
- **Risks or disadvantages.** In this study, there is a possibility that the questions could prompt discussions about difficult relationships or experiences in the workplace.
- **Benefits**. There are no direct benefits in this study. We expect that findings from this study will be used to develop recommendations to inform the design and implementation of quality improvement interventions in neonatal care in Kenya.

**Who is carrying out this study and what is this study about?**

- This study is being carried out by KEMRI in collaboration with the University of Oxford, UK. KEMRI is a Kenyan government organization that carries out medical research to find better ways of preventing and treating illness in the future for everybody’s benefit.
- The study has involved collecting data through non-participant observation from 2 hospitals as well as from health care workers and nurse managers and educational managers of 4 training programmes. Specific for this workshop, we have invited you to contribute to group discussions with other participants and the research team, to make sense of and comment and reflect on the data that has already been collected. You will also help in the co-design of a set of recommendations, as an output from the engagement event. You can leave the event at any time, by letting the researcher know.
- This event will be face-to-face with the requisite Ministry of Health COVID-19 protocols being adhered to (unless we choose to run it online).
- The discussion will be recorded (audio recorded if face-to-face and audio or video recorded if online) to assist later in fully writing up the information.

**Why do you want to talk to me and what does it involve?**

- We have invited you because you have relevant experience in healthcare delivery (particularly neonatal care), research and policy/decision making, and you are familiar with key aspects of how to design recommendations and intervention designs for quality improvement.

**Are there any risks or disadvantages to me for taking part?**

- There are no risks for taking part in the study. However, questions around interactions and social connections can be sensitive as they may include to a discussion about difficult relationships at workplace.
- The discussions should take approximately 5-7 hours. You will be provided with either a standard fare reimbursement of Ksh. 600 or mileage costs at the rate of Ksh.25/km for face-to-face meetings.
- Online stakeholder meeting will be reimbursed with a data voucher worth Ksh.250-500 per person.
- Because we anticipate the meeting to take longer than 3 hours, you will be eligible for a standard out-of-pocket expense of ksh. 1000 per person as determined by KEMRI-WTRP guidelines for study benefits and compensation. This is not meant in any way to influence how you participate in the study.

**Are there any advantages to me for taking part in the study?**

- There are no individual benefits to taking part in this study. In participating in this study, you will contribute in making recommendations in the design of interventions that improve quality of care of neonates in Kenya and elsewhere in the future, for example through developing new health policies.

**Data Protection**

**Who will have access to information about me in this research?**

- All our research records are stored securely in locked cabinets and password protected computers and are accessed only by authorized persons.
- The personally identifiable information you provide will be used only for study purposes such as contact you for scheduling of visits and verification of consent through signed consent forms.
- We will share anonymized individual and summary information we collect or generate with our partner institution, University of Oxford in ways that do not reveal individual participants’ identities.
- Anonymised data will be stored in a secure KEMRI-Wellcome Trust Research Programme repository and may be used for future research.
- KEMRI will keep any keep any personally identifiable information about you from this study for 10 years after the study has finished in accordance with applicable Data Protection Requirements both in Kenya and in the UK. You have the right to access the personal data we hold that pertains to you, to object to or make corrections to the processing of all or part of the personal data. However, this might be limited due to coding which might make unblinding difficult.

**Will the research be published? Could I be identified from any publications or other research outputs?**

- The research will be published. We might however include direct quotations, but without identifying you, in any research outputs.
- In future, information collected or generated during this study may be used to support new research by other researchers in Kenya or other countries on improving newborn service delivery. In all cases, we will only share information with other researchers in ways that do not reveal individual participants’ identities. For example, we will remove such as their names and where they live and replace this information with number codes. Any future research using information from this study must first be approved by a local or national expert committee to make sure that the interests of participants and their communities are protected.
- University of Oxford is responsible for ensuring that staff involved in the study in Kenya adhere to the safe and proper use of any personal information you provide. You can contact the research team for any further information about how your data will be managed. Further information about your rights with respect to your personal data is available from <https://compliance.admin.ox.c.uk/individual-rights>

**Who has allowed this research to take place?**

All research at KEMRI has to be approved before it begins by an independent national committee, an international committee and a committee at the KEMRI Wellcome Trust Research Programme. The committees look carefully at planned work before it begins, and must agree that the research is important, relevant to Kenya and follows nationally and internationally agreed research guidelines. This includes ensuring that all participants’ safety and rights are respected. This study will also seek ethics approval from a subcommittee of the University of Oxford Central University Research Ethics Committee.

**What will happen if I refuse to participate?**

All participation in research is voluntary. You are free to decide if you want to take part or not. If you do agree you can change your mind at any time without any consequences. You can withdraw yourself from the stakeholder event, without giving a reason, and without negative consequences, by advising us of this decision. If you withdraw from the study after the event, information you have already provided will still be included the analysis, however the information you have provided will not be used as quotes.

**What if I have any questions?**

You are free to ask me any question about this research. If you have any further questions about the study, you are free to contact the research team using the contacts below:

**If you want to ask someone independent anything about this research please contact:**

Community Liaison Manager, KEMRI Wellcome Trust Research Programme, P.O. Box 230, Kilifi. Telephone: 041 7522 063, Mobile 0723 342 780 or 0705 154 386

***And***

### KEMRI-Wellcome Trust Research Programme Verbal Assent form for Stakeholder workshop event for: -

**Study title: Pathways Study: Understanding health worker social ties and communication, and their influence in neonatal care.**

**Lay Title: Understanding how health worker interactions influence the care of newborn babies.**

I have followed the study procedure to obtain verbal assent from the participants.

I attest that the information concerning this research was accurately explained to and apparently understood by the participants who have been given opportunity to ask questions which have been answered satisfactorily.

**I confirm that verbal assent was provided (or denied) by the participant(s) listed below who agreed (did not agree) to take part in this research.**

Agree	Refused

Participant Name: _______________________________________________________	□	□

Participant Name: _______________________________________________________	□	□

Participant Name: _______________________________________________________	□	□

Participant Name: _______________________________________________________	□	□

Participant Name: _______________________________________________________	□	□

Participant Name: _______________________________________________________	□	□

Participant Name: _______________________________________________________	□	□

Participant Name: _______________________________________________________	□	□

Participant Name: _______________________________________________________	□	□

Participant Name: _______________________________________________________	□	□

Participant Name: _______________________________________________________	□	□

Participant Name: _______________________________________________________	□	□

Participant Name: _______________________________________________________	□	□

Participant Name: _______________________________________________________	□	□

Participant Name: _______________________________________________________	□	□

Participant Name: _______________________________________________________	□	□

Participant Name: _______________________________________________________	□	□

Participant Name: _______________________________________________________	□	□

Participant Name: _______________________________________________________	□	□

Participant Name: _______________________________________________________	□	□

Participant Name: _______________________________________________________	□	□

Participant Name: _______________________________________________________	□	□

Participant Name: _______________________________________________________	□	□

Participant Name: _______________________________________________________	□	□

**Designee/investigator’s signature:** ____________________________ **Date** ____________

**Designee/investigator’s name:** _____________________________**Time** ____________

(Please print name)
